# Polygenic Resilience Scores are Associated with Lower Penetrance of Schizophrenia Risk Genes, Protection Against Psychiatric and Medical Disorders, and Enhanced Mental Well-Being and Cognition

**DOI:** 10.1101/2024.06.03.24308377

**Authors:** Jonathan L. Hess, Eric J. Barnett, Jiahui Hou, Stephen V. Faraone, Stephen J. Glatt

**Affiliations:** Department of Psychiatry & Behavioral Sciences, Norton College of Medicine at SUNY Upstate Medical University, Syracuse, NY USA; Department of Neuroscience & Physiology, Norton College of Medicine at SUNY Upstate Medical University, Syracuse, NY USA

**Author notes:** **Please Address Correspondence to:** Dr. Stephen J. Glatt 3710 Neuroscience Research Building Institute for Human Performance 505 Irving Avenue Syracuse, NY 13210, Dr. Jonathan Hess 3732 Neuroscience Research Building Institute for Human Performance 505 Irving Avenue Syracuse, NY 13210.

**Keywords:** biobank, bipolar disorder, depression, genome-wide association study, high risk, polygenic score, schizophrenia, suicide, resilience

## Abstract

In the past decade, significant advances have been made in finding genomic risk loci for schizophrenia (SCZ). This, in turn, has enabled the search for SCZ resilience loci that mitigate the impact of SCZ risk genes. Recently, we discovered the first genomic resilience profile for SCZ, completely independent from the established risk loci for SCZ. We posited that these resilience loci protect against SCZ for those having a heighted genomic risk for SCZ. Nevertheless, our understanding of genetic resilience remains limited. It remains unclear whether resilience loci foster protection against adverse states associated with SCZ risk related to clinical, cognitive, and brain-structural phenotypes. To address this knowledge gap, we analyzed data from 487,409 participants from the UK Biobank, and found that resilience loci for SCZ afforded protection against lifetime psychiatric (schizophrenia, bipolar disorder, anxiety, and depression) and non-psychiatric medical disorders (such as asthma, cardiovascular disease, digestive disorders, metabolic disorders, and external causes of morbidity and mortality). Resilience loci also protected against self-harm behaviors, improved fluid intelligence, and larger whole-brain and brain-regional sizes. Overall, this study sheds light on the range of phenotypes that are significantly associated with resilience loci within the general population, revealing distinct patterns separate from those associated with SCZ risk loci. Our findings indicate that resilience loci may offer protection against serious psychiatric and medical outcomes, co-morbidities, and cognitive impairment. Therefore, it is conceivable that resilience loci facilitate adaptive processes linked to improved health and life expectancy.

## INTRODUCTION

Over the past decade, genome-wide association studies (**GWAS**s) have found many loci contributing to risk for schizophrenia (**SCZ**). These efforts led to the development of a polygenic risk score, which accounts for ∼20% of the genetic liability toward SCZ.^1^ The identification of reliable risk markers, in turn, paved the way for studies of the genomic underpinnings of resilience. Resilience buffers against the impact of risk genes for SCZ, which could explain why some individuals are spared from the disorder despite having high levels of genetic liability. Recently, we discovered the first-known genetic resilience profile for SCZ, which comprised thousands of protective alleles at single nucleotide polymorphisms (**SNPs**) enabling some individuals in the population to avoid developing SCZ despite carrying an elevated genetic predisposition.^2^ We also found a significant positive correlation between resilience and risk scores, which was seen in unaffected individuals at high genetic risk for SCZ yet absent in affected individuals with comparable genetic risk. Hence, we posited that, in unaffected individuals, genomic resilience increases with their genomic risk to counteract effects of risk loci, thereby conferring resistance against SCZ.

In this study, we tested the hypothesis that our SCZ resilience score would moderate the impact of SCZ risk on phenotypes and outcomes associated with schizophrenia among 487,409 participants in the UK Biobank. We analyzed extensive genome-wide SNP data in conjunction with lifetime ICD-10 diagnoses gathered from hospital reports, fluid intelligence scores, educational attainment, self-reported mental health outcomes, self-reported lifetime self-harm measures relevant to suicide, and neuroimaging-derived phenotypes of whole-brain and brain-regional sizes. Taken together, our study provides new knowledge about the moderating effects of resilience scores spanning multiple diagnoses and clinically relevant outcomes within a population-representative sample from the UK.

## METHODS

A graphical abstract is provided in **Figure 1** depicting a simplified overview of the current study.

**Figure 1.**
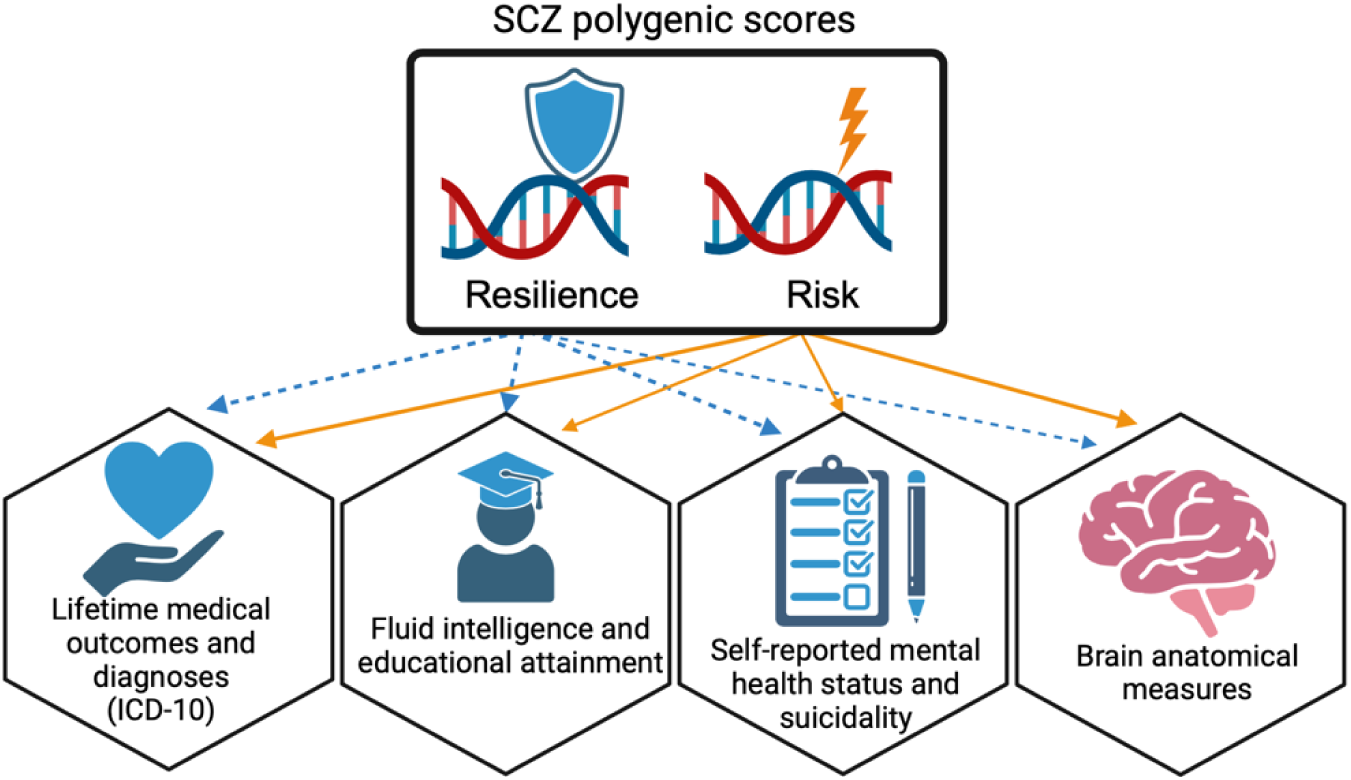
A graphical abstract of the current study. We investigated main and interactive effects of schizophrenia (SCZ) risk and resilience scores on four domains: (1) cognitive performance and reserve measured by fluid intelligence and educational attainment, (2) medical outcomes (includes hospital-recorded ICD-10 codes), (3) self-reported mental health status and measures of self-harm, and (4) neuroimaging-derived phenotypes from structural magnetic resonance imaging (sMRI) scans. Data for this investigation was obtained from the UK Biobank. Images for this figure were generated with BioRender.

### Datasets and Quality Control

We accessed genome-wide genotypes and phenotypic data from 487,409 participants from the UK Biobank. The genome-wide genotypic data were received post-imputation to the Haplotype Reference Consortium (HRC) panel. This was the same reference panel used to impute the genotype datasets from the Schizophrenia Working Group of the Psychiatric Genomics Consortium in which we derived the SNP weights for the SCZ risk and resilience-scoring formulae.^1^ Basic quality control procedures were applied to these data to exclude uncommon variants with a minor allele frequency (MAF) of < 1% and variants found to significantly depart from Hardy-Weinberg equilibrium (*p* ≤ 1e-06). We used the top five genetic principal components released by UK Biobank covering all 487,409 participants (data-field: 22009) throughout our downstream analyses to account for genetic variation due to ancestry.

### Polygenic Scoring

For each participant in the UK Biobank, we used existing formulae to compute genetic scores for SCZ risk and resilience, which represent the weighted sum of risk and resilience-associated effects of SNPs from independent loci. We calculated genetic scores using the software *Plink2*. For risk-scoring, we selected SNPs that had a maximum risk-association *p*-value threshold of 0.05, which was found to maximize the explainable variation in SCZ.^1,2^ Likewise, for resilience-scoring, we selected resilience SNPs with a maximum resilience-association *p*-value threshold of 0.3, which was found to maximize the explainable variation in resilience status.^2,3^ The resilience scores were initially derived to reduce the potential for linkage disequilibrium (LD) between resilience-associated SNPs and risk SNPs for SCZ by solely considering SNPs within low LD regions (*R*^2^<0.2) of risk SNPs. Thus, these resilience scores reflect protective effects from residual genetic variation that remains independent of known risk-related genetic variants for SCZ (including the alternate alleles of risk SNPs), which would theoretically confer protection against SCZ rather than tagging risk effects. For our primary analysis, we focused on resilience scores that were derived using this initial LD pruning threshold of *R*^2^<0.2. However, to ensure that no SNPs of very small risk effects might have unduly influenced our resilience scores, we repeated our analysis using a more stringently defined set of resilience SNPs that exhibit exceptionally low LD with known risk loci, at a threshold of *R*^2^<0.1. We note that it is impractical to filter out variants exhibiting any non-zero LD with known risk loci for SCZ, which would effectively eliminate nearly all variants across the genome and consequently exclude genuine resilience loci. We assessed the similarity of results obtained from our models using the two separate resilience scores.

### Statistical Analyses of Lifetime Medical Diagnoses

We used logistic regression models to estimate the main and interaction effects of resilience and SCZ risk scores on hospital-reported lifetime medical outcomes, diagnoses, and comorbidities recorded in International Classification of Diseases (ICD-10) codes for each UK Biobank participant. To account for the relatively low prevalence of disease outcomes within the UK Biobank, which tended to skew towards older and healthier participants,^4^ we applied a two-pronged approach. Several disorders and conditions of interest to our investigation had exceedingly low numbers of cases (dozens to a low hundreds), hence we applied grouping strategy to increase the number of positive outcomes based on collections of related ICD-10 diagnoses. We referred to these downstream as “broadly defined ICD-10 diagnoses”. Specifically, we aggregated participants who had *any* diagnoses within a particular chapter, referring to those individuals as “cases” and those without any diagnoses within the same chapter as “controls”. An exception was made for ICD-10 Chapter XX (“External causes of morbidity and mortality”), wherein we aggregated diagnoses of participants based on four sub-chapters. These sub-chapters delineate various sources of morbidity or mortality as designated by ICD-10 codes starting with letters “V” (such as transportation-related accidents), “”W” (such as falls or environmental exposures), “X” (such as unspecified exposures, poisoning, intentional self-harm), and “Y” (such as adverse outcomes related to medical care or sequalae of other origin). We analyzed data from a total of 17 ICD-10 chapters and sub-chapters using logistic regression models. We used logistic regression models to estimate the interaction effects of resilience and SCZ risk scores in the presence of their main effects and covariates. We specified the top five principal components as covariates to adjust for genetic variation due to ancestry.

Additionally, we examined select ICD-10 codings that contained a minimum of 1,000 cases, using logistic regression models to estimate the main and interaction effects of resilience and SCZ risk scores. We refer to these downstream as “narrowly defined ICD-10 diagnoses”. This analysis included neuropsychiatric disorders (schizophrenia, bipolar disorder, depression, anxiety, substance abuse), neurodegenerative diseases (Alzheimer’s disease, vascular dementia, and unspecified dementia), chronic inflammatory or immunologic conditions (Crohn’s disease, asthma, primary biliary cirrhosis [also known as “primary biliary cholangitis” or “fibrosis and cirrhosis of the liver”]), and metabolic conditions (type I and type II diabetes mellitus). These conditions were selected for analysis as they have been observed to have significant genetic overlap with SCZ.^5–8^ In each of the above-described analyses, we adjusted for multiple comparisons using the Benjamini-Hochberg false-discovery rate (FDR)-adjustment procedure. This same significance criteria was applied to all subsequent statistical analyses.

### Statistical Analysis of Mental Health Questionnaires

To complement our analyses of ICD-10 clinical codes, we explored moderating effects of resilience loci in the context of mental health self-reports. Our objective in integrating mental health self-assessments into our study was to capture potential links between resilience loci and mental or behavioral outcomes that may hold clinical relevance, even if those outcomes do not meet criteria for a formal psychiatric diagnosis. We analyzed data obtained from a mental health questionnaire offered to UK Biobank participants upon enrollment, yielding a total of 19 items of mental health status completed by 486,974 participants.^9^ Using logistic regression models, we estimated the main and interaction effects of resilience and SCZ risk scores on the likelihood of respondents answering “yes” to each questionnaire item. Covariates included in our regression models were the top five genome-wide PCs to adjust for genetic variation due to ancestry, participants’ age at recruitment, and participants’ sex.

### Statistical Analyses of Fluid Intelligence and Educational Attainment

We hypothesized that resilience loci might enhance fluid intelligence, which is often impaired among individuals affected with SCZ.^10–12^ Fluid intelligence was assessed among UK Biobank participants at enrollment (field ID: 20016, *n*=160,002) with each score representing a simple unweighted sum of the total number of correct answers from 13 fluid intelligence questions. We employed multiple linear regression models to estimate the main and interaction effects of resilience and SCZ risk scores on inverse-rank normalized scores for fluid intelligence scores while accounting for ancestry PCs, together with age at assessment and sex of participants.

Furthermore, we hypothesized that resilience loci might be associated with greater educational attainment. Exploring this hypothesis holds importance as SCZ is known to be associated with lower educational attainment.^13^ Moreover, greater education is associated with increased levels of cognitive reserve, which protects against cognitive impairment later in life,^14–16^ and reduces risk for mortality.^17^ Educational qualifications (field ID: 6138) were collected from UK Biobank participants at the time of enrollment (*n*=480,714). Following the approach described by Ge *et al.*^18^ we mapped qualification onto one of seven categories defined by the 1997 International Standard Classification of Education (ISCED) of the United Nations Educational, Scientific, and Cultural Organization. We employed multiple linear regression models to estimate the main and interaction effects of resilience and SCZ risk scores on inverse-rank normalized scores for educational qualifications while accounting for ancestry PCs, age at assessment, and sex.

### Statistical Analysis of Self-Reported Suicidality

SCZ is associated with greater lifetime risk for self-harm, suicide behavior, and suicide attempt than the general population.^19–21^ Therefore, we hypothesized that resilience loci might moderate the penetrance of SCZ risk genes, protecting against suicidality. To test this hypothesis, we analyzed two variables linked to suicidality corresponding to self-reported lifetime measures of self-harm ideation or engagement (*i.e.,* “Ever contemplated self-harm” [data field 20485] and “Ever self-harmed” [data field 20480]). These variables were obtained from an online mental health questionnaire completed by UK Biobank participants at the time of enrollment, of which 157,297 participants responded. We estimated both the main and interaction effects of resilience and SCZ risk on the likelihood of respondents endorsing each of these self-harm measures. We specified the top five genome-wide PCs to adjust for genetic variation due to ancestry, participants’ age at recruitment, and participants’ sex as covariates in each model.

### Statistical Analysis of Structural Brain Phenotypes

Prior studies have observed widespread reductions in cortical and sub-cortical brain region sizes among individuals with SCZ.^22,23^ In light of these results, we performed an exploratory analysis to test the moderating effects of resilience loci on processed imaging derived phenotypes (**IDPs**) representing structural profiles for 240 distinct brain parcellations. The IDPs were collected at a single time-point among four imaging centers in the UK (*i.e.,* Cheadle, Reading, Newcastle upon Tyne, or Bristol) from the year 2014 onward. Standardized cortical and subcortical atlases were used to extract left and right hemispheric measures of cortical surface area, volume, and thickness, and subcortical regional brain volumes. Brain-regional phenotypes were standardized to a mean of 0 and variance of 1 prior to statistical analysis. We used linear regression models to estimate both the main and interaction effects of resilience and SCZ risk scores on brain-regional sizes. The covariates included in our regression models were: (1) the top five genotype-based principal components to adjust for variation in ancestry, (2) self-reported sex, (3) age at brain-imaging assessment, (4) site of imaging assessment, (5) weight, height, and body mass index (BMI) at time of imaging assessment, (6) a volumetric scaling factor for T1-head image to standard space (data-field 25000), and (7) variation in location of the scanner and head position described in three-dimensional coordinates (data-fields 25756, 25757, 25758).^24^ Main effects of resilience and risk were also evaluated in separate models that excluded their interaction effect.

## RESULTS

Across all 300 outcome measures that we analyzed, we observed strong, significant agreement between the main effects of the two resilience scores using LD *R*^2^ threshold of 0.2 and 0.1 (Pearson’s *r* = 0.90, *p* = 1.37×10^-108^) (**Supplementary Figure 1**). Henceforth, we focus on the results from our models that used resilience scores calculated using the LD *R*^2^ threshold of 0.2, which had better statistical power.

Resilience scores showed an opposite direction of effect than SCZ risk scores for 193 of the outcomes examined. Furthermore, the main effects of resilience and SCZ risk scores were significantly negatively correlated across all outcomes (Pearson’s *r* = -0.47, *p* = 3.2×10^-18^) (**Supplementary Figure 2**). These findings lend support to our hypothesis that resilience scores moderate the penetrance of SCZ risk genes.

All the regression models in our study included the top five genome-wide PCs as covariates to adjust for potential confounding relationships due to genetic variation between ancestries. Our primary rationale for including PCs as covariates in our models was to adjust for possible admixture and population stratification, which could artificially inflate the effects and increase the number of false-positives. We observed widespread, significant associations between ancestry-related PCs and the outcome measures included in our study (**Supplementary Figures 2 – 8)**.

### Resilience Scores Moderate Effects of SCZ Risk on Narrowly Defined ICD-10 Diagnoses

We found seven narrowly defined ICD-10 diagnoses wherein resilience scores interacted significantly with SCZ risk scores, reducing the likelihood of diagnosis (FDR*p*<0.05) (**Figure 2, Supplementary File 1**). Significant interaction effects were associated with protection against the following diagnoses: bipolar disorder (BD) (interaction: OR = 0.93, 95% CI 0.89 – 0.98, *z*-score = -2.7, FDR*p* = 0.017), depressive episodes (interaction: OR = 0.96, 95% CI 0.95 – 0.98, *z*-value = -4.64, FDR*p* = 1.710^-5^), anxiety disorder (interaction: OR = 0.97, 95% CI 0.95 – 0.99, *z*-value = -6.1, FDR*p* = 6.410^-3^), tobacco abuse (interaction: OR = 0.94, 95% CI 0.93 – 0.95, *z*-value = -8.1, FDR*p* = 1.410^-14^), and alcohol abuse (interaction: OR = 0.93, 95% CI 0.90 – 0.96, *z*-value = -5.1, FDR*p* = 2.910^-6^). In addition, we replicated our previously reported findings that resilience scores reduce the penetrance of SCZ risk loci, reducing the likelihood of SCZ (interaction: OR = 0.93, 95% CI 0.90 – 0.97, *z*-value = -3.32, FDR*p* = 3.510^-3^). These findings suggest that resilience scores moderate the penetrance of SCZ risk loci, providing *cross-protection* against multiple psychiatric outcomes related to SCZ.

**Figure 2.**
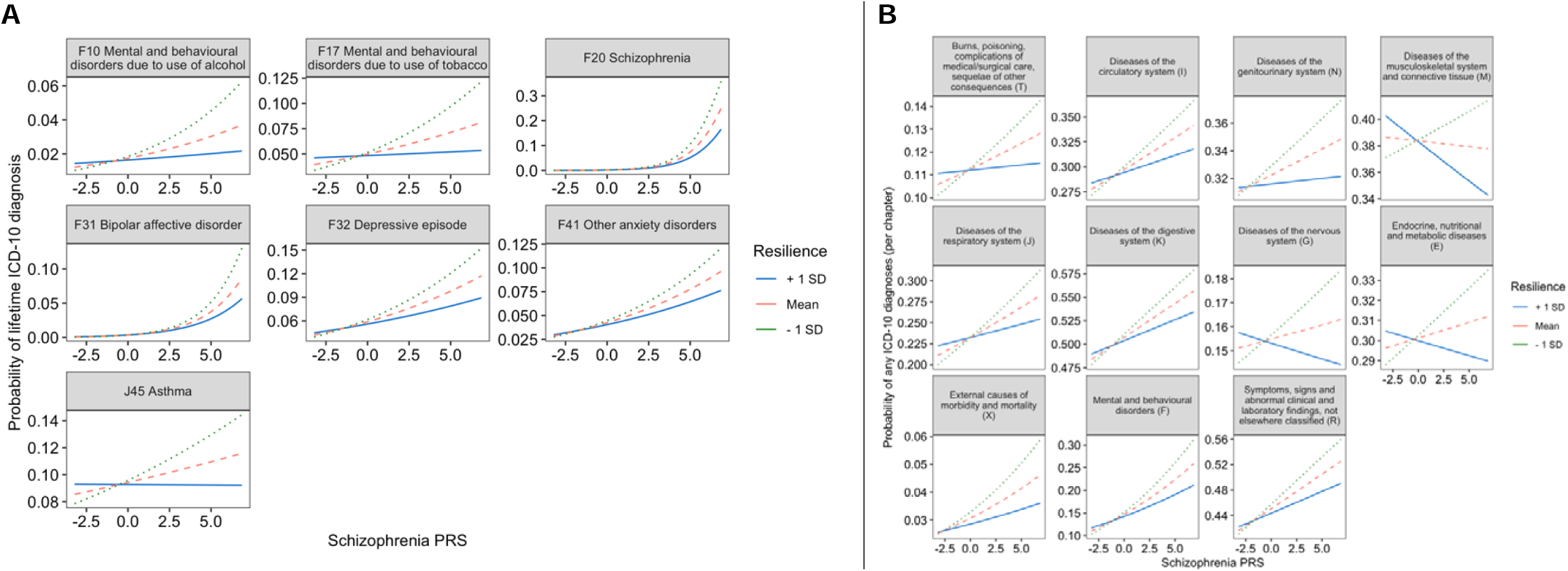
Line graphs depicting statistically significant interactive effects of schizophrenia (SCZ) risk and resilience scores on hospital-reported time diagnoses among UK Biobank participants spanning neuropsychiatric, immunologic, and metabolic disorders based on the 10^th^ revision of the International Classification of Diseases (ICD). All records were obtained from UK Biobank participants at the time of enrollment and reflect lifetime diagnostic codes. The results shown in this plot for resilience scores correspond to the scores calculated using resilience-associated SNPs that exhibit minimal linkage disequilibrium with mild-risk SNPs for SCZ (*R*^2^ value of 0.2 or less). (**A**) Line graphs depicting significant interaction effects of resilience and SCZ risk scores on narrowly defined ICD-10 diagnoses. The y-axes show the fitted probabilities of the ICD-10 diagnoses. The solid blue line denotes the effect of SCZ risk among individuals with a resilience score one standard deviation above the sample mean. The dashed light-red line denotes the effect of SCZ risk at a mean level of resilience. The dotted green line denotes the effect of SCZ risk among individuals with a resilience score one standard deviation below the sample mean. (**B**) Line graph depicting the significant interactions of resilience and SCZ risk scores on probability of *any* ICD-10 diagnoses *per* chapter. The y-axes show the probabilities of chapter-specific ICD-10 diagnoses among UK Biobank participants. The x-axes represents the standardized SCZ risk scores. Each line represents the effect of SCZ risk scores on likelihood of participants having any ICD-10 diagnoses based on levels of resilience. The solid blue line depicts the effect of SCZ risk scores among individuals with resilience one standard deviation above the sample mean. The dashed light-red line depicts the effect of SCZ risk scores at mean levels of resilience. The dotted green line depicts the effect of SCZ risk scores among individuals with resilience scores one standard deviation below the sample mean. The lines in the plot depict the effects of SCZ risk at different levels of resilience. We note that y-axes for all plots were scaled according to each outcome variable to enhance visualization of the interaction effects.

Additionally, we found a significant interaction between SCZ risk and resilience scores, associated with protection against asthma (interaction: OR = 0.97, 95% CI 0.96 – 0.98, *z*-value = -6.1, FDR*p* = 1.2810^-8^).

### Resilience Scores Moderate Effects of SCZ Risk on Broadly Defined ICD-10 Diagnoses

As presented in **Figure 2B**, we found significant interaction effects between resilience and SCZ risk scores influencing risk for 11 categories of hospital-reported medical diagnoses, grouped by ICD-10 chapters/sub-chapters. All summary statistics from our regression models are provided in **Supplementary File 1**. High levels of resilience scores attenuated the penetrance of SCZ risk genes, conferring protection against the following categories of diseases and/or co-morbid outcomes:

- *any* mental and behavioral disorders (ICD-10 codes from Chapter V with prefix “F”) (interaction: OR = 0.97, 95% CI 0.96 – 0.98, *z*-value = -6.12, FDR*p* = 3.1×10^-10^)
- *any* disease of the genitourinary system (ICD-10 codes from Chapter XIV with prefix “N”) (interaction: OR = 0.98, 95% CI 0.98 – 0.99, *z*-value = -3.8, FDR*p* = 8.1×10^-4^),
- *any* diseases of the musculoskeletal system and connective tissue (ICD-10 codes from Chapter XIII with prefix “M”) (interaction: OR = 0.97, 95% CI 0.97 – 0.99, *z*-value = -5.7, FDR*p* = 3.1×10^-8^),
- *any* diseases of the respiratory system (ICD-10 codes from Chapter X with prefix “J”) (interaction: OR = 0.979, 95% CI 0.97 – 0.987, *z*-value = -5.1, FDR*p* = 2.2×10^-6^),
- *any* diseases of the nervous system (ICD-10 codes from Chapter VI with prefix “G”) (interaction: OR = 0.98, 95% CI 0.97 – 0.99, *z*-value = -3.4, FDR*p* = 2.4×10^-4^),
- *any* endocrine, nutritional and metabolic diseases (ICD-10 codes from Chapter IV with prefix “E”) (interaction: OR = 0.985, 95% CI 0.98 – 0.99, *z*-value = -3.4, FDR*p* = 3.8×10^-4^),
- *any* disease of the digestive system (ICD-10 codes from Chapter XI with prefix “K”) ( interaction: OR = 0.989, 95% CI 0.98 – 0.995, *z*-value = -3.22, FDR*p* = 0.0025),
- *any* diseases of the circulatory system (ICD-10 codes from Chapter IX with prefix “I”) (interaction: OR = 0.986, 95% CI 0.98 – 0.99, *z*-value = -2.96, FDR*p* = 0.0016)
- *any* diagnoses broadly defined as symptoms, signs, and abnormal clinical and laboratory findings, not elsewhere classified (ICD-10 codes from Chapter XVII with prefix “R”) (interaction: OR = 0.98, 95% CI 0.977 – 0.99, *z*-value = -4.5, FDR*p* = 2.2×10^-5^),
- *any* external causes of morbidity and mortality (ICD-10 codes from Chapter XX with prefix “X”) (interaction: OR = 0.97, 95% CI 0.95 – 0.99, *z*-value = -2.63, FDR*p* = 0.019),
- *any* instances of burns, exposures, complications from medical or surgical care, or sequelae of other consequences (ICD-10 codes from Chapter XIX with prefix “T”) (interaction: OR = 0.98, 95% CI 0.97 – 0.99, *z*-value = -3.11, FDR*p* = 0.0031).

### Resilience Promotes Greater Educational Attainment and Fluid Intelligence

We found that high levels of resilience scores moderated the penetrance of SCZ risk genes, promoting higher fluid intelligence scores (interaction: β = 0.023, *t*-value = 9.17, FDR*p* = 4.7×10^-20^) and greater educational attainment (interaction: β = 0.017, *t*-value = 11.9, FDR*p* = 1.9×10^-32^) (**Figure 3, Supplementary File 1**).

**Figure 3.**
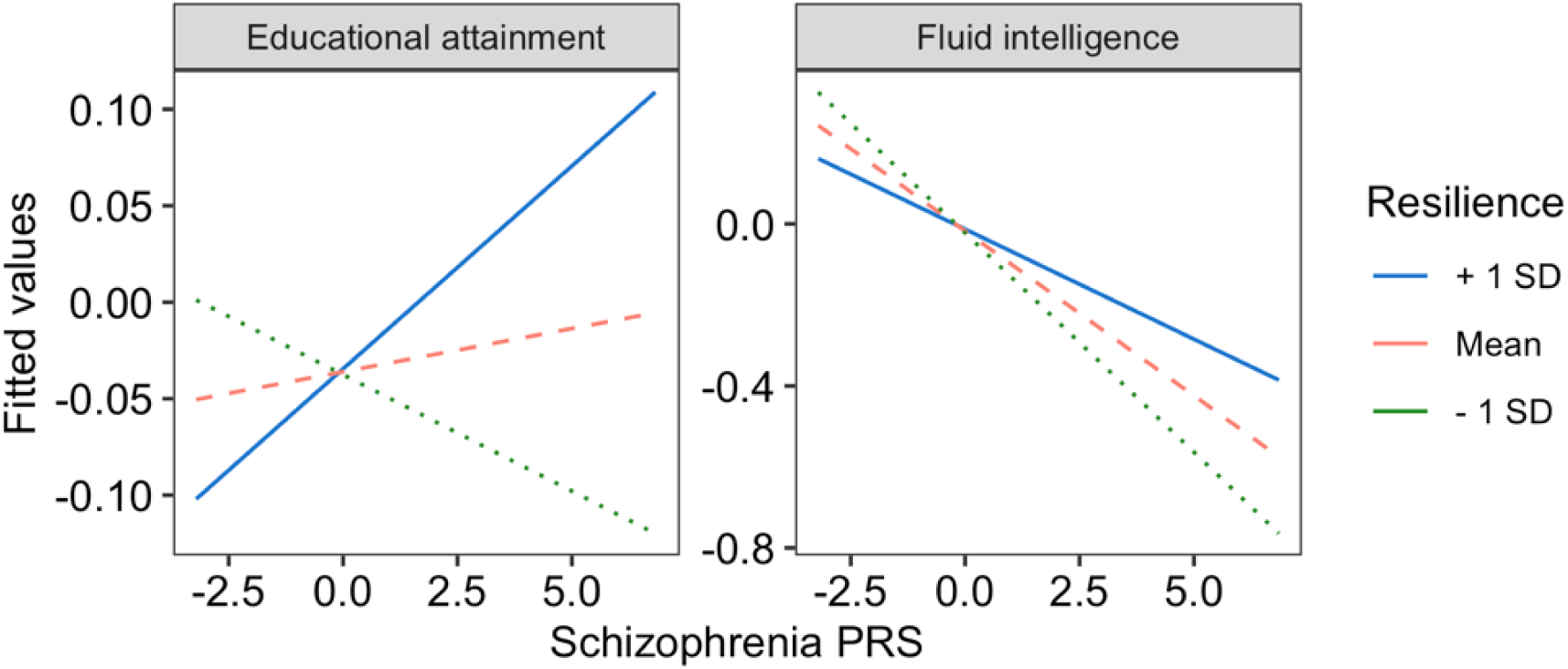
Line graphs depicting statistically significant interactive effects of schizophrenia (SCZ) risk and resilience scores educational attainment and fluid intelligence scores obtained from UK Biobank participants at the time of enrollment. The results shown in this plot for resilience scores correspond to the scores calculated using resilience-associated SNPs that exhibit minimal linkage disequilibrium with mild-risk SNPs for SCZ (*R*^2^ value of 0.2 or less). The y-axes show the fitted probability values for each outcome measure. The lines in the plot depict the effects of SCZ risk at different levels of resilience. The solid blue line denotes the effect of SCZ risk among individuals with a resilience score one standard deviation above the sample mean. The dashed light-red line denotes the effect of SCZ risk at a mean level of resilience. The dotted green line denotes the effect of SCZ risk among individuals with a resilience score one standard deviation below the sample mean. We note that y-axes for all plots were scaled according to each outcome variable to enhance visualization of the interaction effects.

### Resilience Attenuates Risk for Self-Reported Mental Well-Being

We found that resilience scores significantly interact with SCZ risk scores, fostering enhanced well-being for 12 mental health items spanning anxiety, mania, and depressive-related measures, such as: feeling highly irritable or argumentative (interaction: OR = 0.97, *z*-value = -3.8, FDR*p* = 2.9×10^-4^), being seen by a psychiatrist for nerves, tension, and/or depression (interaction: OR = 0.96, *z*-value = -6.9, FDR*p* = 3.4×10^-11^), being seen by a doctor for nerves, tension, and/or depression (interaction: OR = 0.97, *z*-value = -7.3, FDR*p* = 3.6×10^-12^), as well as feeling unenthusiastic or disinterested for a whole week (interaction: OR = 0.98, *z*-value = -4.1, FDR*p* = 1.2×10^-4^), to name a few (**Figure 4A, Supplementary File 1**).

**Figure 4.**
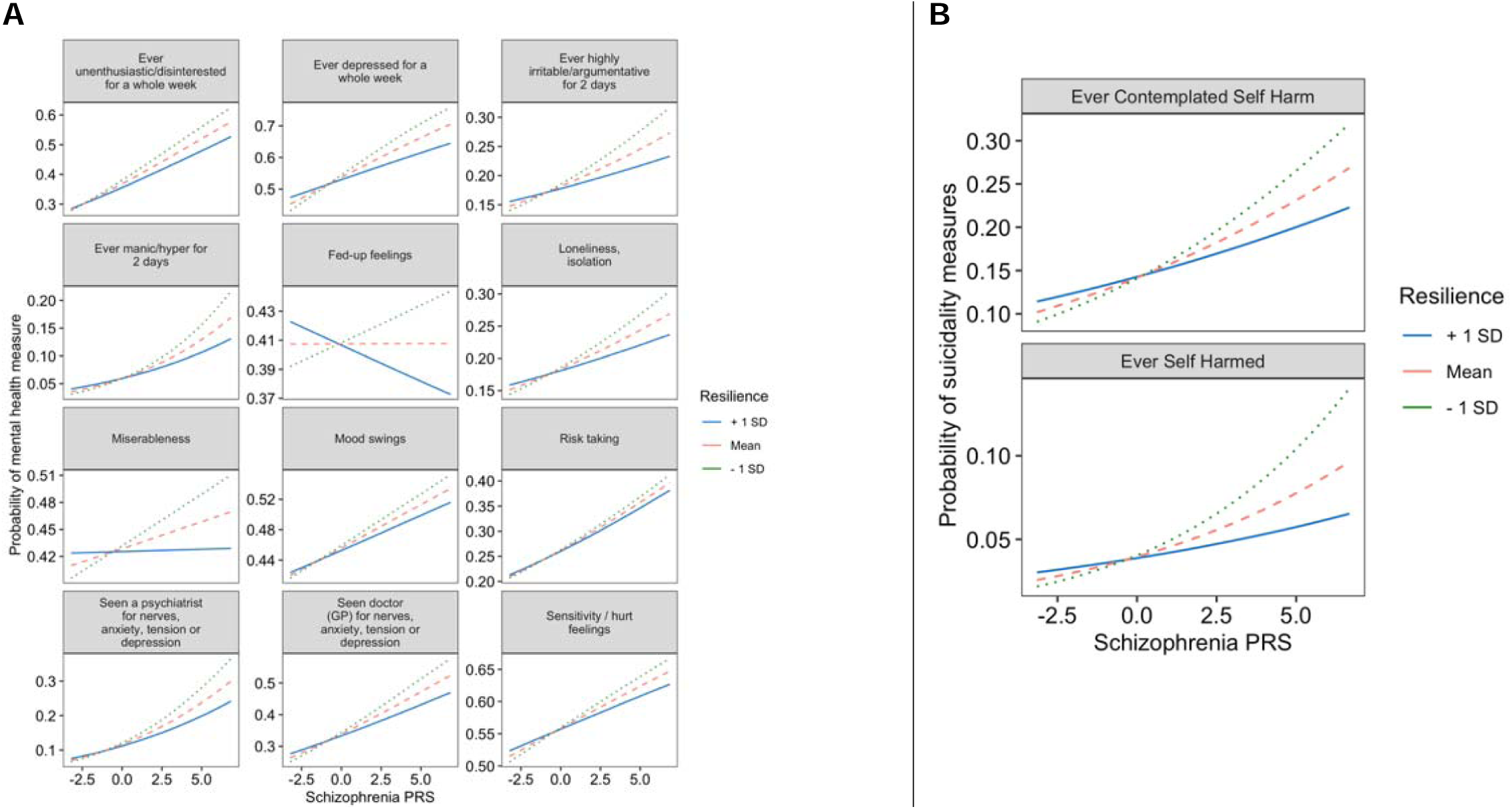
Resilience scores moderate the penetrance of schizophrenia risk scores, enhancing self-reported mental well-being and reducing self-harm behaviors. A graphical representation of statistically interactive effects of schizophrenia (SCZ) risk and resilience scores on self-reported indicators of mental health provided by UK Biobank participants at the time of enrollment. All measures included in our analysis were scored as binary (yes/no) events. The results shown in this plot for resilience scores correspond to the scores calculated using resilience-associated SNPs that exhibit minimal linkage disequilibrium with mild-risk SNPs for SCZ (*R*^2^ value of 0.2 or less). (**A**) Line plots depicting the significant interaction effects of resilience scores and SCZ risk scores on self-reported mental health measures. The x-axes represent the standardized SCZ risk scores. The y-axis shows the fitted probability values of self-reported mental health measures for which resilience and SCZ risk scores exhibited a significant interaction effect. The lines in the plot depict the effects of SCZ risk at different levels of resilience. The solid blue line denotes the effect of SCZ risk among individuals with a resilience score one standard deviation above the sample mean. The dashed light-red line denotes the effect of SCZ risk at a mean level of resilience. The dotted green line denotes the effect of SCZ risk among individuals with a resilience score one standard deviation below the sample mean. (**B**) Line plots depicting the significant interactions of resilience and SCZ risk scores on probability of self-reported suicidality measures. The y-axes show the probabilities of both lifetime engagement in self-harm and ideation of self-harm among UK Biobank participants. The x-axes represent the standardized SCZ risk scores. Each line represents the effect of SCZ risk scores on likelihood of participants any suicidality based on levels of resilience. The solid blue line depicts the effect of SCZ risk scores among individuals with resilience one standard deviation above the sample mean. The dashed light-red line depicts the effect of SCZ risk scores at mean levels of resilience. The dotted green line depicts the effect of SCZ risk scores among individuals with resilience scores one standard deviation below the sample mean. We note that y-axes for all plots were scaled according to each outcome variable to enhance visualization of the interaction effects.

In addition, our findings revealed mental well-being items for which resilience scores demonstrated a significant main effect (FDR*p*<0.05) conferring protection against the following: feeling tense or “high strung” (OR = 0.95, *z*-value = -3.9, FDR*p* = 4.2×10^-4^), being a worrier or holding feelings of anxiousness (OR = 0.97, *z*-value = -3.51, FDR*p* = 1.7×10^-3^), and feelings of nervousness (OR = 0.97, *z*-value = -2.7, FDR*p* = 0.022). However, the interaction effects between resilience and SCZ risk scores for these outcomes were not significant (**Supplementary Figure 9, Supplementary File 1**).

Resilience and SCZ risk scores showed interaction effects associated with protection against self-harm ideation (interaction: OR = 0.95, *z*-value = -4.4, FDR*p* = 2.4×10^-5^) and engagement (interaction: OR = 0.93, *z*-value = -3.8, FDR*p* = 1.3×10^-4^) (**Figure 4B, Supplementary File 1**).

### Resilience Scores Promote Larger Whole-Brain and Brain-Regional Brain Sizes

No significant interaction effects between resilience scores and SCZ risk scores were seen for brain-imaging phenotypes (**Supplementary File 1**). However, we found 15 brain imaging-derived phenotypes for which significant main effects were observed solely for resilience scores, with SCZ risk scores showing non-significant effects. High levels of resilience were found to be associated with increased size of multiple brains regions, including: total grey matter volume of the brain, volume of the cortex, volume and thickness of the fusiform gyrus, bi-hemispheric thickness of the pars opercularis, bi-hemispheric thickness of the rostral middle frontal gyrus, thickness of the superior frontal gyrus, volume and thickness of the medial orbitofrontal gyrus, and thickness of the pars orbitalis (**Figure 5A, Supplementary File 1**).

**Figure 5.**
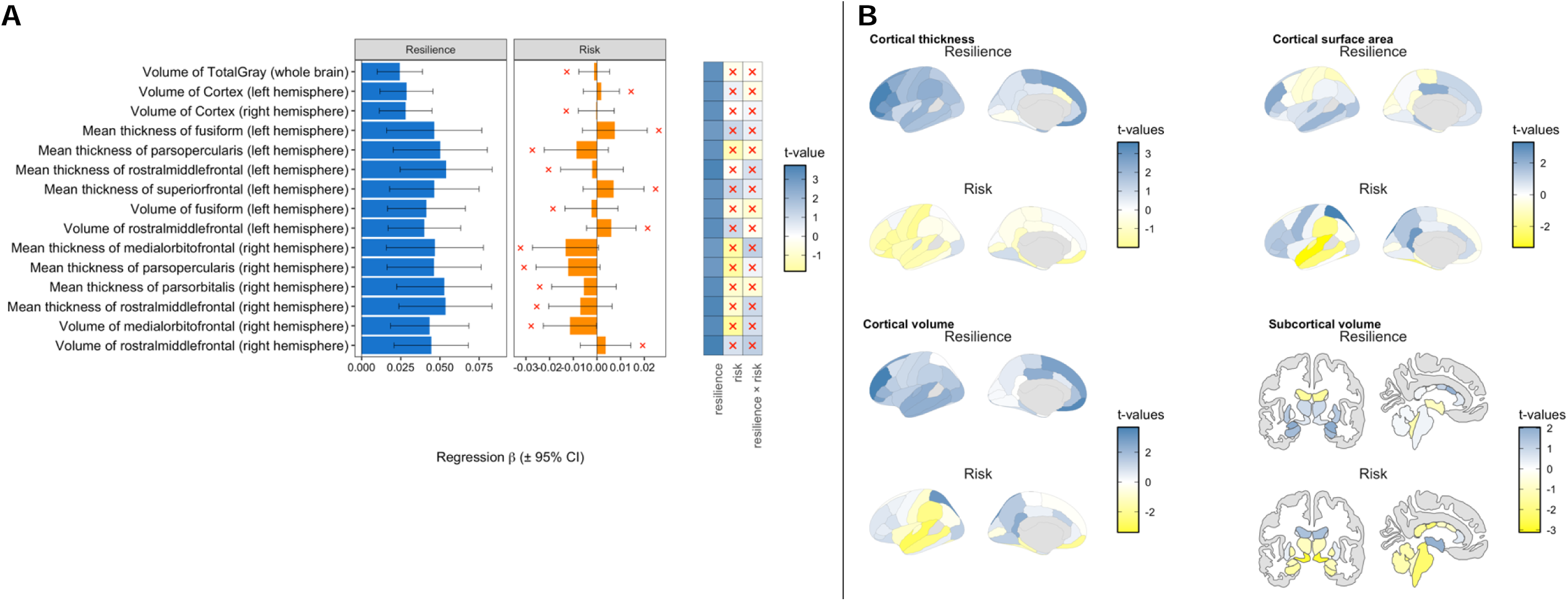
A graphical representation of main and interactive effects of schizophrenia (SCZ) risk and resilience scores on brain imaging-derived phenotypes collected from structural magnetic resonance imaging (sMRI) of UK Biobank participants. The results shown are for sMRI phenotypes that exhibited a significant association with the main effect of resilience scores. The results shown in this plot for resilience scores correspond to the scores calculated using resilience-associated SNPs that exhibit a maximum linkage disequilibrium with mild-risk SNPs for SCZ at a *R*^2^ value of 0.2. (**A**) Bar-plots showing the main effect of resilience and SCZ risk scores on brain-imaging-derived phenotypes for which a significant main effect of resilience was observed. No significant interaction effect of resilience and SCZ risk was observed for any sMRI phenotype, nor were any significant associations observed for the main effect of SCZ risk scores. The bar-plot on the left depicts the main effects of resilience (blue bars) and SCZ risk scores (orange bars), excluding their interactive effect, for each of the 15 corresponding brain imaging-derived phenotypes (y-axis). The main effect of resilience and SCZ risk is presented in terms of the Odds Ratio on the x-axis. The 95% confidence intervals of the Odds Ratio are shown as error bars in the bar-plot. Non-significant main effects (FDR*p* > 0.05) are denoted by a red “x”. The heat-map shown on the right side of the plot presents the significance (FDR*-*adjusted *p*-values, -log_10_ transformed) of the main effects of resilience and SCZ risk scores in the presence of their interactive effect. Similar to the bar-plot, non-significant main or interactive effects are denoted by a red “x”. (**B**) Anatomical brain plots showing the main effects of resilience and SCZ risk scores for brain-regional sizes for all 240 distinct cortical and subcortical parcellations. Regions are color coded according to the magnitude and direction of main effects for SCZ resilience and risk scores, with darker blue indicating a stronger positive association (*i.e*., higher scores associated with larger regional sizes) and brighter yellow indicating a stronger negative association (*i.e*., higher scores associated with smaller regional sizes).

**Figure 5B** provides an anatomical representation of the main effects of resilience and SCZ risk scores, illustrating that higher resilience scores were associated with widespread increases in cortical brain thickness, regional increases in surface area particularly within the frontal and temporal lobe, and increases in subcortical volume. Conversely, high SCZ risk scores were associated with widespread reductions in cortical and subcortical brain-regional sizes.

## DISCUSSION

In prior studies, we posited that genomic loci associated with resilience moderate the influence of risk genes for SCZ, thereby protecting “resilient” individuals who have a high risk for schizophrenia. Our current study aimed to improve our understanding of genetic resilience for SCZ by investigating the main effect of resilience scores, as well as interactive effects of resilience and SCZ risk scores, across multiple genetic and phenotypic scales, including: (1) lifetime medical diagnoses, (2) fluid intelligence and educational attainment, (3) self-reported mental well-being outcomes and measures of suicidality, and (4) anatomical measures of whole brain and brain-regional size.

Our results showed that resilience scores exhibit significant interactions with SCZ risk scores, buffering the impact of SCZ risk genes on a range of outcomes including cognitive indicators, psychological states, and psychiatric and medical outcomes. It is remarkable that for all the interaction plots we see the quintessential signature of resilience. They show resilience protecting those at high risk for outcomes but having no effect at lower levels of risk.

From our analyses that focused primarily on ICD-10 diagnoses as outcome measures, we found that resilience scores significantly attenuated the impact of SCZ risk genes, reducing the likelihood of psychotic (schizophrenia), mood (BD and depression), substance abuse (tobacco and alcohol), and anxiety disorders among UK Biobank participants. Collectively, this is the first-ever reported evidence of genetic *resilience* profiles for SCZ exhibiting *bona fide* cross-protection across multiple lifetime neuropsychiatric outcomes.

In light of these results, it is plausible that genetic *resilience* architectures might mirror the pattern of shared protective genes among neuropsychiatric disorders.^25^ Initially, resilience scores for SCZ were derived without accounting for co-morbidities or pleotropic effects. Hence, there is potential to expand and refine the SCZ resilience scores to encompass additional cross-protective effects. Such insights might only emerge through joint genome-wide association studies of multiple psychiatric disorders. Refinements to our resilience scores, such as integrating additional samples into our analyses to capture risk genes with very small effect sizes and incorporating cases with other psychiatric disorders, may lead to improved detection of moderating effects of resilience scores on shared pleotropic risk genes. Another avenue that is still unexplored is the possibility of disorder-specific resilience factors. Prior studies have identified disorder-specific genetic risk factors for SCZ and BD.^26^ Hence, it is plausible that an analogous yet undiscovered pattern of disorder-specificity exists for resilience loci. Further investigations are needed to explore resilience loci that may differ across psychiatric disorders.

Multiple studies have shown that SCZ is associated with lower rates of educational attainment, lower intelligence scores, and cognitive impairment.^13,27,28^ This suggests that SCZ risk genes may influence cognitive functioning and cognitive reserve,^29,30^ leading to impairment in cognitive performance and poorer clinical outcomes. Our study showed that resilience scores moderated the penetrance of SCZ risk loci, with their interaction effect promoting greater rates of educational attainment, and higher fluid intelligence scores.

We found that resilience profiles moderated the penetrance of SCZ risk loci for 12 items from the mental well-being questionnaire within. Higher resilience scores protected against numerous emotional, psychological, and behavior-related outcomes such as miserableness, loneliness, risk taking, mania, prolonged episodes of anhedonia, and prolonged depressive symptoms, and depressive symptoms severe enough to prompt a healthcare visit, to name a few. These findings lend support to the hypothesis that high levels of resilience loci may foster enhanced states of psychological resilience, improved mental well-being, and better adaptation to adversity.

Increased genetic vulnerability for SCZ was significantly associated with increased risk for ideation and engagement in self-harm behaviors, in agreement with the well-documented relationship between SCZ and increased risk for suicide attempt, self-harm, and suicide ideation.^19–21^ While resilience scores did not exhibit a significant main effect on either suicidality measure, our results demonstrated a significant interaction effect between resilience and SCZ risk. The interaction revealed that high resilience scores offset the adverse effects of SCZ risk, reducing the probability of individuals engaging in or contemplating self-harm. The effect sizes of the interaction term on these self-harm measures indicated that for each unit increase in both risk and resilience scores, the likelihood of self-harm contemplation or engagement decreased by 5–7%, respectively. This finding holds implications for suicide and psychiatric research. Yet, the number of respondents to the question about lifetime ideation or engagement in self-harm represented about one-third of the total number of participants in the UK Biobank, thus these findings will need to be evaluated in independent cohorts. The rate of attempted suicide is substantially higher among individuals with a pre-existing psychiatric disorder. Furthermore, suicide ranks among the leading causes of death globally, with rates of suicides steadily increasing over the past two decades.^31^ Identifying risk factors for suicide serves a critical step in accurately predicting those at elevated risk and in improving intervention and prevention. Moreover, it remains to be shown whether resilience loci foster protective effects against suicidality independent from the interaction effect with SCZ risk genes.

Due to selection biases, the UK Biobank is skewed towards older, healthier participants.^4^ Therefore, our study lacked sufficient statistical power to detect significant associations between resilience scores and individual ICD-10 codes, particularly those outcomes that are already relatively uncommon in the general population, but further skewed to an even rarer frequency within the UK Biobank, such as schizophrenia. Hence, our analytic strategy examined *any* instances of a condition within individual ICD-10 chapters, with one exception being the ICD-10 Chapter XX wherein we aggregated sub-items of a chapter, to enhance the number of positive outcomes for more cohesively defined external causes of co-morbidity and mortality. This came at a cost of granularity, as we lacked statistical power to link resilience scores to individual diseases or conditions. Yet, our results of diagnostic groups provided several important insights into types of pathology, as well as causes of morbidity and mortality against which resilience scores show protective effects. Our analysis of lifetime medical diagnoses stored in hospital-reported ICD-10 codes for UK Biobank participants showed that resilience scores significantly attenuated the probability of individuals being diagnosed with any diseases among two ICD-10 chapters that are chiefly relevant to brain disorders, namely: *any* mental or behavioral disorder, and *any* nervous system disorder. In addition, we discovered that resilience scores moderate the effect of SCZ risk scores, reducing the overall probability of participants being diagnosed with conditions not specifically originating in central nervous system, namely: cardiovascular diseases, disease of musculoskeletal or connective tissues, disease of the genitourinary system, diseases of the digestive tract, as well as diseases of the endocrine and metabolic system. Moreover, we found that resilience scores moderated the effect of SCZ risk, reducing the probability of individuals succumbing to external causes of morbidity or mortality, which included instances of accidents, injury, assaults, self-harm, medical or surgical complications, or sequalae of other causes. These findings provide insights on the clinical relevance of resilience scores within a population-level sample. Moreover, our results show that moderating effects of resilience scores extend beyond psychiatric and mental health outcomes. Our findings suggest that resilience loci might install a general capacity to adapt to various adverse conditions, including reducing risk for diseases that manifest in peripheral tissues and organ systems beyond the central nervous system.

A multitude of epidemiological studies show that SCZ is associated with risk for numerous serious medical co-morbidities of relevance to our key findings, including urinary infections^32,33^ and sexually transmitted infections,^34^ chronic kidney disease,^35^ gastrointestinal disease,^36,37^ cardiovascular disease,^38,39^ and metabolic disorders such as diabetes.^40^ Metabolic dysfunction is a major clinical concern in SCZ as patients are prone to poor dietary habits, unhealthy lifestyles (such as increased smoking behavior and inactivity), and metabolic alterations resulting from use of second-generation or atypical antipsychotic medications. Epidemiological data also highlights SCZ as a leading causes of morbidities and mortality.^41^ Taken together, our results demonstrate that resilience scores reduce probability of serious medical outcomes and co-morbidities that have been firmly linked with SCZ in prior work. This underscores the potential clinical and epidemiological significance of our results, as resilience loci may foster protection against adverse outcomes associated with reduced quality of life, increased disability, and premature mortality.

Our study also found significant associations between resilience scores and increased total grey matter and size of multiple cortical brain regions. However, no significant association was observed between brain structure and SCZ genetic liability, nor did we observe a significant interactive effect between resilience and SCZ risk in the context of brain structure. Less than 10% of UK Biobank participants have been currently assessed *via* structural brain-imaging, thus posing a potential limitation on the power to detect statistically significant effects. Regardless, the main effect of resilience scores on increased total grey matter and brain-regional sizes of the cortex hints at potential protective mechanisms against SCZ genetic liability. Furthermore, resilience and SCZ risk scores exhibited distinct patterns of effects on brain structure. Higher resilience scores were associated with widespread increases in thickness and volume of the cortex, as well as selective increases in cortical surface area particularly in the frontal and temporal lobes, and selective increases in subcortical brain volumes. Conversely, SCZ risk scores were associated with a distinct profile of cortical reductions in thickness, as well as selective reductions in cortical surface area and volume, as well as selective reductions in subcortical brain volumes. While many of the brain-region-specific effects of resilience and SCZ risk scores did not reach statistical significance, their directions of effect on brain structure seem to be clearly divergent. In accordance, the Enhancing Neuro Imaging Genetics through Meta-Analysis (ENIGMA) Consortium found that individuals with SCZ, on average, show widespread reductions in cortical thickness, with multiple brain regions being affected. On average, cortical surface area is shown to be globally reduced in SCZ, with regional differences in surface area appearing comparatively small. Our findings suggest that resilience loci might foster an adaptive increase in cortical brain size to compensate for adverse effects of SCZ risk genes. However, these results are based on single time-point, cross-sectional measures of regional brain sizes. Prospective follow-up measures of brain size are anticipated in the UK Biobank, which could offer insights into the role of resilience loci in adaptive remodeling of brain structure. Our key brain-structural findings indicated that resilience scores were associated with increases in cortical thickness and volume in multiple regions of the frontal lobe, in addition to increased volume and thickness of the ventral temporal lobe, particularly the fusiform gyrus. Notably, previous evidence shows that individuals at ultra-high risk for psychosis present with significant reductions in the fusiform gyrus, as well as regions of the frontal lobe that we found to increase in size commensurate with resilience scores.^22,42,43^ The link we found between the fusiform gyrus and resilience scores mirrors an association initially reported by Hettwer *et al*. who found a positive correlation between resilience scores and increased volume and area of the fusiform gyrus in a sample of 101 research participants with no personal or family history of mental illness.^44^

It is important to acknowledge that our study did not account for individual differences in lifestyle or environmental factors, which could very well have a significant influence on the results we presented. Furthermore, the interactions that we modeled were limited to genetic proxies based on common variants, thus influences of rare variants are not reflected in our analyses. Hence, future studies should investigate the possibility of gene-by–environment interactions in the context of risk and resilience, aiming to elucidate moderating effects of genetic variants, both common and rare, in conjunction with non-genetic factors.

We have previously demonstrated significant and replicable protective effects of resilience scores against SCZ in multiple independent datasets.^2,3^ The results from our current study further strengthen this evidence, serving as an additional external validation. However, it is important to acknowledge that potential selection biases and population admixture in the UK Biobank may have influenced our findings. The accuracy of genetic scores can be affected by differences in linkage disequilibrium (LD) structure between the base population in which the genetic scores were initially derived and the populations in which the scores were computed. While the UK Biobank is predominantly composed of individuals of European descent, there are a mix of non-European individuals, which could potentially lead to biases in genetic findings. We explicitly adjusted for ancestry with the use of genome-wide PCs as covariates to mitigate their potential confounding effects. Yet, there remains the possibility of unaccounted-for confounding due to higher-order interactions between ancestry variation and the calculated genetic scores. Ancestry variation demonstrated significant associations with a multitude of phenotypic measures examined in our study, encompassing ICD-10 codes, self-reported mental health outcomes, and brain-structural profiles. This suggests that genetic diversity across ancestries can result in notable phenotypic variations, as evidenced by patterns in the UK Biobank dataset. Genetic ancestry PCs were used as covariates throughout all our analyses to account for these confounding effects. Furthermore, genetic scores for resilience and SCZ risk merely reflect the influences of heritable variation and do not capture non-genetic components of phenotypic variability. environmental factors were not integrated into our models. We adjusted for participant age at recruitment and sex to address potential variation in phenotypic outcomes stemming from demographic differences. Additionally, we included domain-specific covariates for our analyses of the structural brain-imaging-derived phenotypes to mitigate ascertainment biases due to site-level variation and measurement noise due to head motion during image acquisition. These represent conservative steps aimed at ensuring the robustness of our primary results against likely sources of confounding. However, it remains possible that cryptic sources of ascertainment bias may have influenced our findings. As such, an important next step toward confirming the reliability of our results is conducting multiple rounds of replication in external datasets, including from other biobanks. Despite these technical considerations, population-based samples hold distinct advantages over genetic datasets that are acquired principally through clinical ascertainment. For example, the genetic relationships identified in the UK Biobank are expected to reflect signals present in the population as a whole, in contrast to relationships observed among a subgroup of patients that may be highly homogenous in terms of their clinical presentation, thus limiting statistical power to discover genetic effects relevant at the population-level. Altogether, a crucial next step in advancing this line of research is to assess the reproducibility of our findings, especially in context of population-based studies with compositional characteristics different from the UK Biobank cohort.

Taken together, this study provides us with key insights into genetic and phenotypic links with resilience loci for SCZ. We observed a multitude of outcomes for which resilience loci moderated the influence of SCZ risk genes. Overall, our findings demonstrate first-ever evidence that resilience loci foster cross-protective effects against psychiatric and immunologic genetic risk profiles, as well as evidence that resilience scores attenuate rates of serious medical outcomes and co-morbidities, including lowering rates of suicidality measures. Utilizing phenome-wide information from the UK Biobank offers considerable value for our understanding of resilience loci. However, biobank-level datasets are currently limited in scale with lower-level molecular profiles. Embracing multi-scale ‘omic approaches that encompass profiles such as transcriptome-wide profiles of transcribed gene products (including non-coding or protein-coding RNAs), proteome-wide levels of proteins, lipidomic, and metabolomic profiles, may offer crucial insights into the biological and functional relevance of resilience loci within tissues relevant to pathophysiology. Collectively, our results suggest that individuals high genomic resilience scores have a have reduced risk for psychiatric and serious medical outcomes and co-morbidities, many of which are associated with reduced life expectancy. As such, we postulate that resilience loci might facilitate adaptation to adverse outcomes, potentially enhancing longevity. Nonetheless, further research is needed to explore this hypothesis. If the results from our study are substantiated and extended to biological and environmental measures, we anticipate significant advancements in resilience genetics offering invaluable insights for future development of improved diagnostics, interventions, and prevention strategies against disease.

## Data Availability

This research has been conducted using the UK Biobank Resource under Application Number 60050. Custom-written scripts used for analysis are available upon request made to the corresponding author, but release of individual-level data is not permitted.

https://www.ukbiobank.ac.uk/

## ACKNOWLEDGEMENTS

Dr. Hess is supported by grants from the U.S. National Institute of Mental Health (R21MH126494), the U.S. National Institute of Neurological Disorders and Stroke (R01NS128535), the Central New York Community Fund, and NARSAD: The Brain & Behavior Research Foundation. Dr. Faraone’s research is supported by the European Union’s Horizon 2020 research and innovation programme under grant agreement 965381; NIH/NIMH grants U01AR076092-01A1, R0MH116037, 5R01AG064955-02, 1R21MH126494-01, 1R01NS128535-01, R01MH131685-01, 1R01MH130899-01A1, Corium Pharmaceuticals, Tris Pharmaceuticals and Supernus Pharmaceutical Company. Dr. Glatt is supported by grants from the U.S. National Institutes of Health (R01MH101519, R01AG054002, R01AG064955, and R21MH126494), the Sidney R. Baer, Jr. Foundation, and NARSAD: The Brain & Behavior Research Foundation.

## REFERENCES

1. Ripke, S. et al. Biological insights from 108 schizophrenia-associated genetic loci. Nature 511, 421–427 (2014).

2. Hess, J. et al. A polygenic resilience score moderates the genetic risk for schizophrenia. Mol. Psychiatry (2019).

3. Hess, J. L. et al. A polygenic resilience score moderates the genetic risk for schizophrenia: Replication in 18,090 cases and 28,114 controls from the Psychiatric Genomics Consortium. Am. J. Med. Genet. B Neuropsychiatr. Genet. e32957 (2023) doi:10.1002/ajmg.b.32957.

4. Fry, A. et al. Comparison of Sociodemographic and Health-Related Characteristics of UK Biobank Participants With Those of the General Population. Am. J. Epidemiol. 186, 1026– 1034 (2017).

5. Tylee, D. S. et al. An Atlas of Genetic Correlations and Genetically Informed Associations Linking Psychiatric and Immune-Related Phenotypes. JAMA Psychiatry 79, 667–676 (2022).

6. Tylee, D. S., et al. Genetic Correlations among Brain-Behavioral and Immune-Related Phenotypes Based on Genome-Wide Association Data. 070730–070730 http://biorxiv.org/lookup/doi/10.1101/070730 (2016).

7. Chen, J. et al. Genetic Relationship between Alzheimer’s Disease and Schizophrenia. Alzheimers. Dement. 18, e065861 (2022).

8. DeMichele-Sweet, M. a. A., et al. Genetic risk for schizophrenia and psychosis in Alzheimer disease. Mol. Psychiatry 23, 963–972 (2018).

9. Biobank, U. K. UK Biobank Mental health web-based questionnaire (Version 1.3) Resource 22. https://biobank.ctsu.ox.ac.uk/crystal/refer.cgi?id=22.

10. Martin, A. K., Mowry, B., Reutens, D. & Robinson, G. A. Executive functioning in schizophrenia: Unique and shared variance with measures of fluid intelligence. Brain Cogn. 99, 57–67 (2015).

11. Roca, M. et al. The relationship between executive functions and fluid intelligence in schizophrenia. Front. Behav. Neurosci. 8, 46 (2014).

12. Keyes, K. M., Platt, J., Kaufman, A. S. & McLaughlin, K. A. Association of Fluid Intelligence and Psychiatric Disorders in a Population-Representative Sample of US Adolescents. JAMA Psychiatry 74, 179–188 (2017).

13. Tesli, M. et al. Educational attainment and mortality in schizophrenia. Acta Psychiatr. Scand. 145, 481–493 (2022-5).

14. Wilson, R. S. et al. Education and cognitive reserve in old age. Neurology 92, e1041– e1050 (2019).

15. Zahodne, L. B., Stern, Y. & Manly, J. J. Differing effects of education on cognitive decline in diverse elders with low versus high educational attainment. Neuropsychology 29, 649–657 (2015-7).

16. Clouston, S. A. P. et al. Education and Cognitive Decline: An Integrative Analysis of Global Longitudinal Studies of Cognitive Aging. J. Gerontol. B Psychol. Sci. Soc. Sci. 75, e151– e160 (2020).

17. Balaj, M. et al. Effects of education on adult mortality: a global systematic review and meta-analysis. The Lancet Public Health 9, e155–e165 (2024).

18. Ge, T. et al. The Shared Genetic Basis of Educational Attainment and Cerebral Cortical Morphology. Cereb. Cortex 29, 3471–3481 (2019).

19. Pompili, M. et al. Suicide risk in schizophrenia: learning from the past to change the future. Ann. Gen. Psychiatry 6, 10 (2007).

20. Bai, W. et al. Worldwide prevalence of suicidal ideation and suicide plan among people with schizophrenia: a meta-analysis and systematic review of epidemiological surveys. Transl. Psychiatry 11, 552 (2021).

21. Jakhar, K., Beniwal, R. P., Bhatia, T. & Deshpande, S. N. Self-harm and suicide attempts in Schizophrenia. Asian J. Psychiatr. 30, 102–106 (2017).

22. van Erp, T. G. M. et al. Cortical Brain Abnormalities in 4474 Individuals With Schizophrenia and 5098 Control Subjects via the Enhancing Neuro Imaging Genetics Through Meta Analysis (ENIGMA) Consortium. Biol. Psychiatry (2018) doi:10.1016/j.biopsych.2018.04.023.

23. Van Erp, T. G. M. et al. Subcortical brain volume abnormalities in 2028 individuals with schizophrenia and 2540 healthy controls via the ENIGMA consortium. Mol. Psychiatry (2016) doi:10.1038/mp.2015.63.

24. UK Biobank: Primary brain imaging documentation. https://biobank.ctsu.ox.ac.uk/crystal/refer.cgi?id=1977.

25. Lee, P. H. et al. Genomic Relationships, Novel Loci, and Pleiotropic Mechanisms across Eight Psychiatric Disorders. Cell (2019) doi:10.1016/j.cell.2019.11.020.

26. Ruderfer, D. M. et al. Genomic Dissection of Bipolar Disorder and Schizophrenia, Including 28 Subphenotypes. Cell 173, 1705–1715.e16 (2018).

27. Ohi, K. et al. A Brief Assessment of Intelligence Decline in Schizophrenia As Represented by the Difference between Current and Premorbid Intellectual Quotient. Front. Psychiatry 8, 293 (2017).

28. McCutcheon, R. A., Keefe, R. S. E. & McGuire, P. K. Cognitive impairment in schizophrenia: aetiology, pathophysiology, and treatment. Mol. Psychiatry 28, 1902–1918 (2023).

29. Herrero, P. et al. Influence of cognitive reserve in schizophrenia: A systematic review. Neurosci. Biobehav. Rev. 108, 149–159 (2020).

30. Barnett, J. H., Salmond, C. H., Jones, P. B. & Sahakian, B. J. Cognitive reserve in neuropsychiatry. Psychol. Med. 36, 1053–1064 (2006).

31. World Health Organization. Suicide worldwide in 2019. https://www.who.int/publications-detail-redirect/9789240026643.

32. Graham, K. L., Carson, C. M., Ezeoke, A., Buckley, P. F. & Miller, B. J. Urinary tract infections in acute psychosis. J. Clin. Psychiatry 75, 379–385 (2014).

33. Miller, B. J. et al. A prevalence study of urinary tract infections in acute relapse of schizophrenia. J. Clin. Psychiatry 74, 271–277 (2013).

34. Liang, C.-S. et al. The Risk of Sexually Transmitted Infections Following First-Episode Schizophrenia Among Adolescents and Young Adults: A Cohort Study of 220 545 Subjects. Schizophr. Bull. 46, 795–803 (2020).

35. Tzeng, N.-S. et al. Is schizophrenia associated with an increased risk of chronic kidney disease? A nationwide matched-cohort study. BMJ Open 5, e006777 (2015).

36. Severance, E. G., Yolken, R. H. & Eaton, W. W. Autoimmune diseases, gastrointestinal disorders and the microbiome in schizophrenia: more than a gut feeling. Schizophr. Res. 176, 23–35 (2016).

37. Severance, E. G., Prandovszky, E., Castiglione, J. & Yolken, R. H. Gastroenterology issues in schizophrenia: why the gut matters. Curr. Psychiatry Rep. 17, 27 (2015-5).

38. Fan, Z., Wu, Y., Shen, J., Ji, T. & Zhan, R. Schizophrenia and the risk of cardiovascular diseases: A meta-analysis of thirteen cohort studies. J. Psychiatr. Res. 47, 1549–1556 (2013).

39. Hennekens, C. H., Hennekens, A. R., Hollar, D. & Casey, D. E. Schizophrenia and increased risks of cardiovascular disease. Am. Heart J. 150, 1115–1121 (2005).

40. Suvisaari, J., Keinänen, J., Eskelinen, S. & Mantere, O. Diabetes and Schizophrenia. Curr. Diab. Rep. 16, 16 (2016).

41. Correll, C. U. et al. Mortality in people with schizophrenia: a systematic review and meta-analysis of relative risk and aggravating or attenuating factors. World Psychiatry 21, 248–271 (2022-6).

42. Jung, S. et al. Fusiform gyrus volume reduction associated with impaired facial expressed emotion recognition and emotional intensity recognition in patients with schizophrenia spectrum psychosis. Psychiatry research. Neuroimaging 307, (2021).

43. Lee, C. U. et al. Fusiform Gyrus Volume Reduction in First-Episode Schizophrenia: A Magnetic Resonance Imaging Study. Arch. Gen. Psychiatry 59, 775–781 (2002).

44. Hettwer, M. D. et al. Evidence From Imaging Resilience Genetics for a Protective Mechanism Against Schizophrenia in the Ventral Visual Pathway. Schizophr. Bull. 48, 551– 562 (2022).

