## Supplementary Figure 1 - 9 for "Polygenic Resilience Scores are Associated with Lower Penetrance of Schizophrenia Risk Genes, Protection Against Psychiatric and Medical Disorders, and Enhanced Mental Well-Being and Cognition"

**Supplementary Figures**

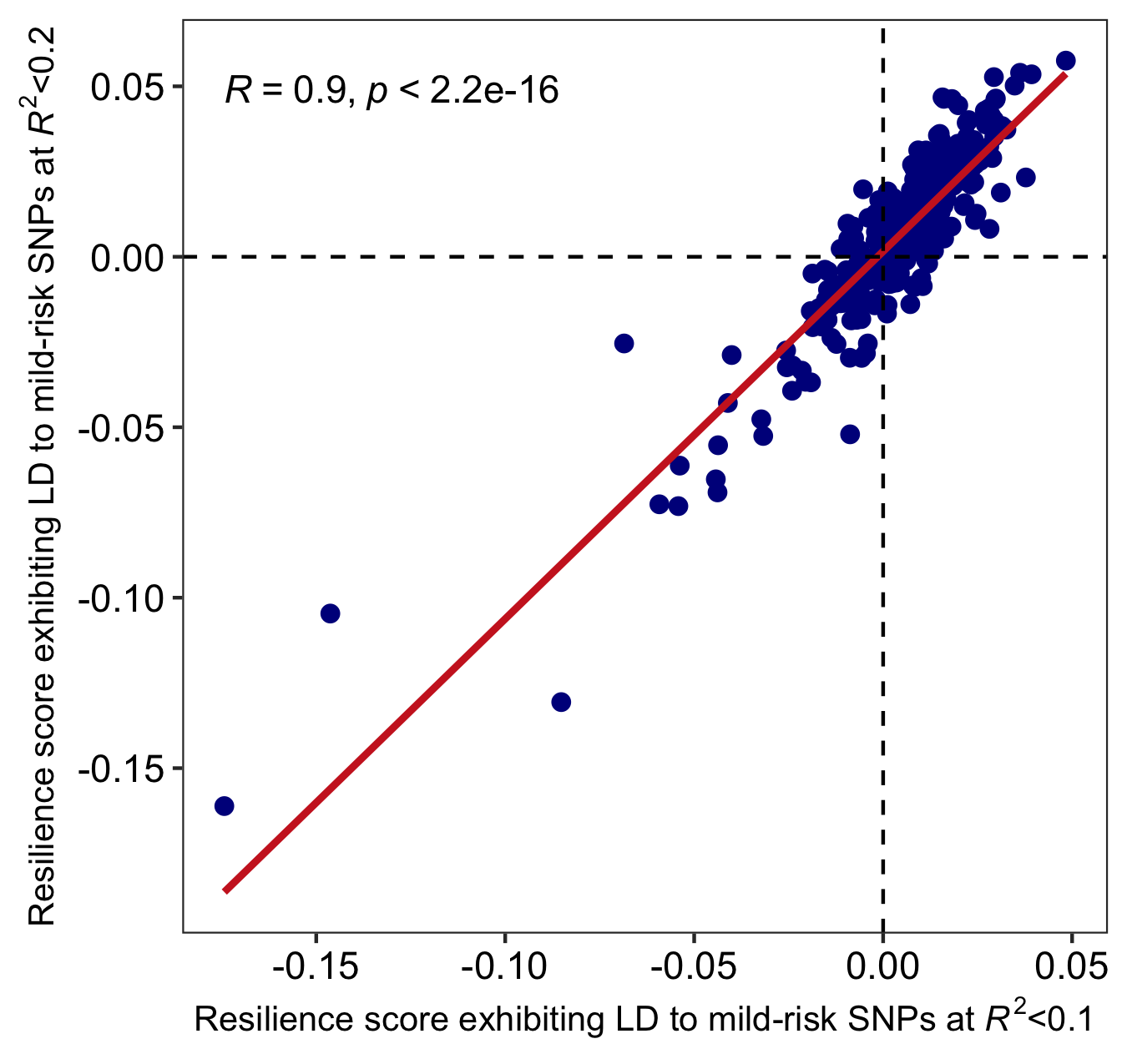

**Supplementary Figure 1.** Scatterplot depicting the agreement in main effects for resilience scores calculated in the UK Biobank using two different resilience-associated SNPs that were pruned with different values to reduce linkage disequilibrium (LD) to known risk SNPs for SCZ. The x-axis corresponds to scores derived using the more stringent LD pruning threshold of *R*^2^=0.1 (indicating that resilience-associated SNPs exhibited LD with risk SNPs at a *R*^2^ value less than 0.1. The y-axis corresponds to scores derived using the less stringent LD pruning threshold of *R*^2^=0.2, indicating that resilience-associated SNPs had a maximum LD with risk SNPs at a *R*^2^ value less than 0.2. Each dot corresponds to each of the 280 distinct outcome measures that were examined in our study, including: (1) educational attainment and fluid intelligence, (2) lifetime medical diagnoses at enrollment based on hospital reported ICD-10 codes, (3) self-reported mental health status, (4) self-reported self-harm measures, and (5) brain imaging-derived phenotypes. The two resilience scores exhibited a strong and significant similarity of their main effects on outcome measures.

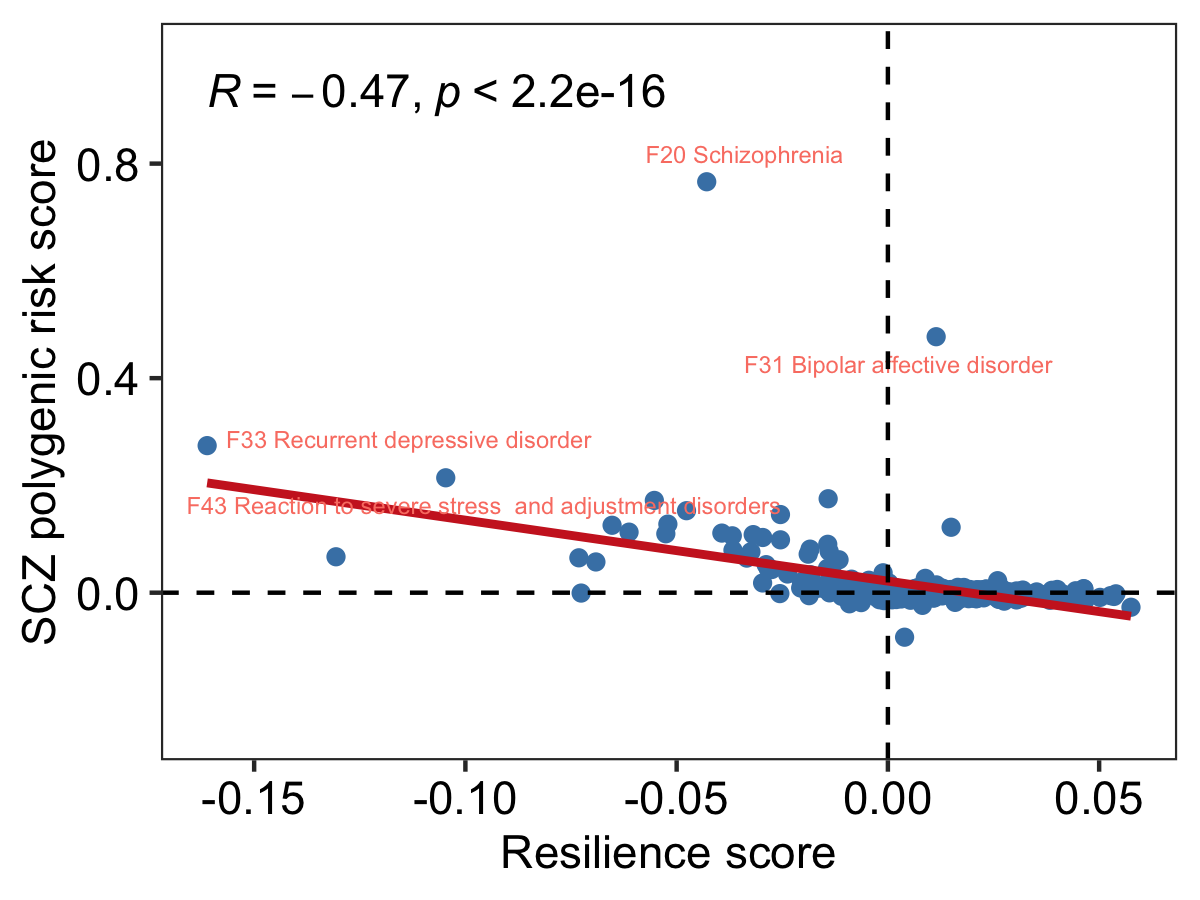

**Supplementary Figure 2.** A scatterplot showing the relationship between main effects of resilience and SCZ risk scores on across all outcome measures investigated in this study. A significant negative correlation was observed, as indicated in the plot. The solid red line designates the best-fit linear relationship between main effects of resilience and SCZ risk scores.

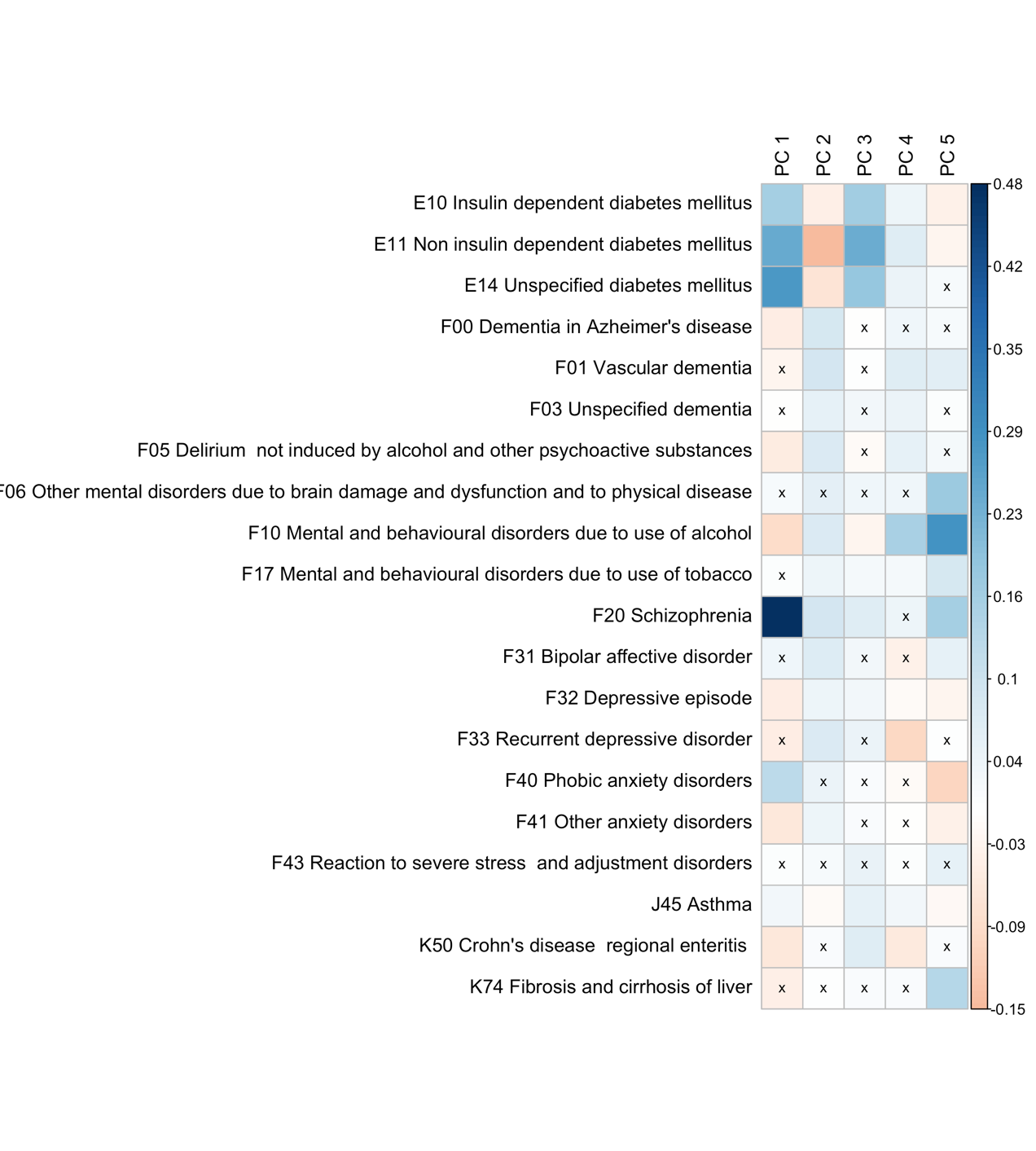

**Supplementary Figure 3.** Heatmap depicting the magnitude and significance of correlations between ICD-10 diagnoses for neuropsychiatric, immunologic, and metabolic disorders (y-axis) and the top five genome-wide principal components (PCs) related to ancestry (x-axis) in the UK Biobank. The color of each box represented the magnitude and direction of the Pearson’s *r* correlation coefficient. Boxes with no cross mark (“x”) represent nominally significant associations between genetic risk scores and genome-wide PCs.

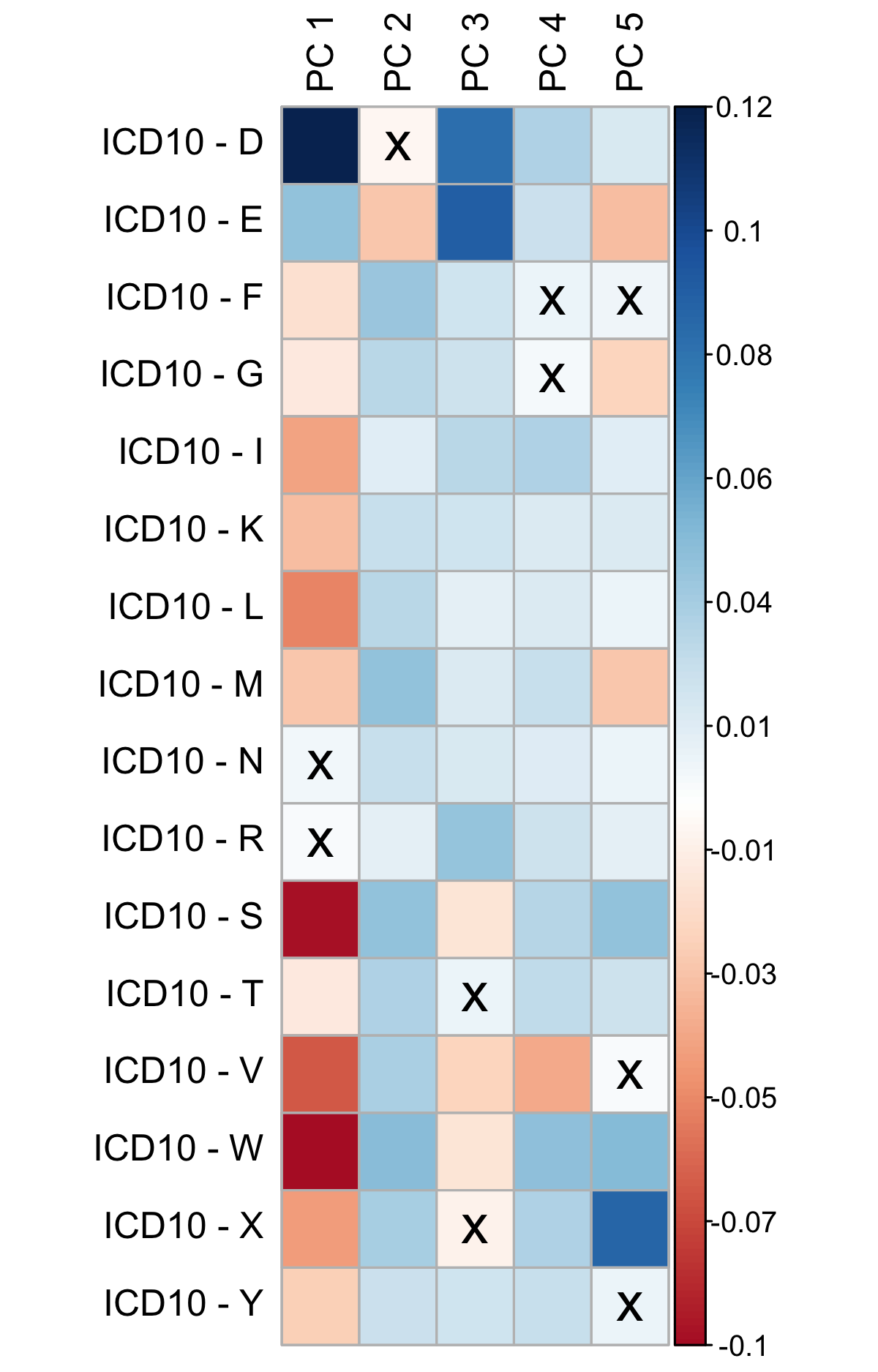

**Supplementary Figure 4.** Heatmap depicting the magnitude and significance of correlations between lifetime diagnoses based on International Classification of Diseases 10^th^ revision grouped by chapter (y-axis) and the top five genome-wide principal components (PCs) related to ancestry (x-axis) in the UK Biobank. The color of each box represented the magnitude and direction of the Pearson’s *r* correlation coefficient. Boxes with no cross mark (“x”) represent nominally significant associations between ICD10 diagnoses and genome-wide PCs.

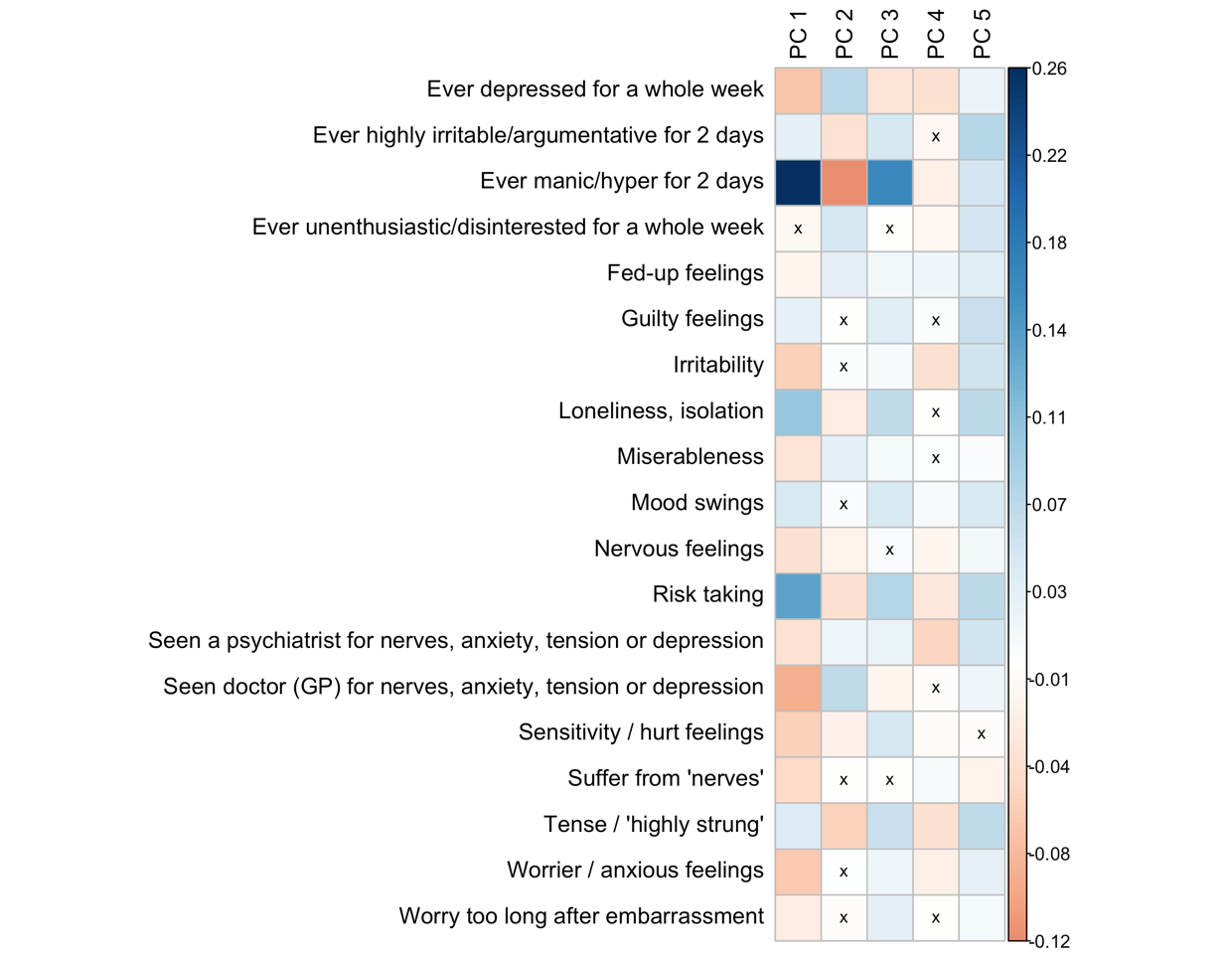

**Supplementary Figure 5.** Heatmap depicting the magnitude and significance of correlations between items from a self-reported mental well-being questionnaire (y-axis) and the top five genome-wide principal components (PCs) related to ancestry (x-axis) in the UK Biobank. The color of each box represented the magnitude and direction of the Pearson’s *r* correlation coefficient. Boxes with no cross mark (“x”) represent nominally significant associations between ICD10 diagnoses and genome-wide PCs.

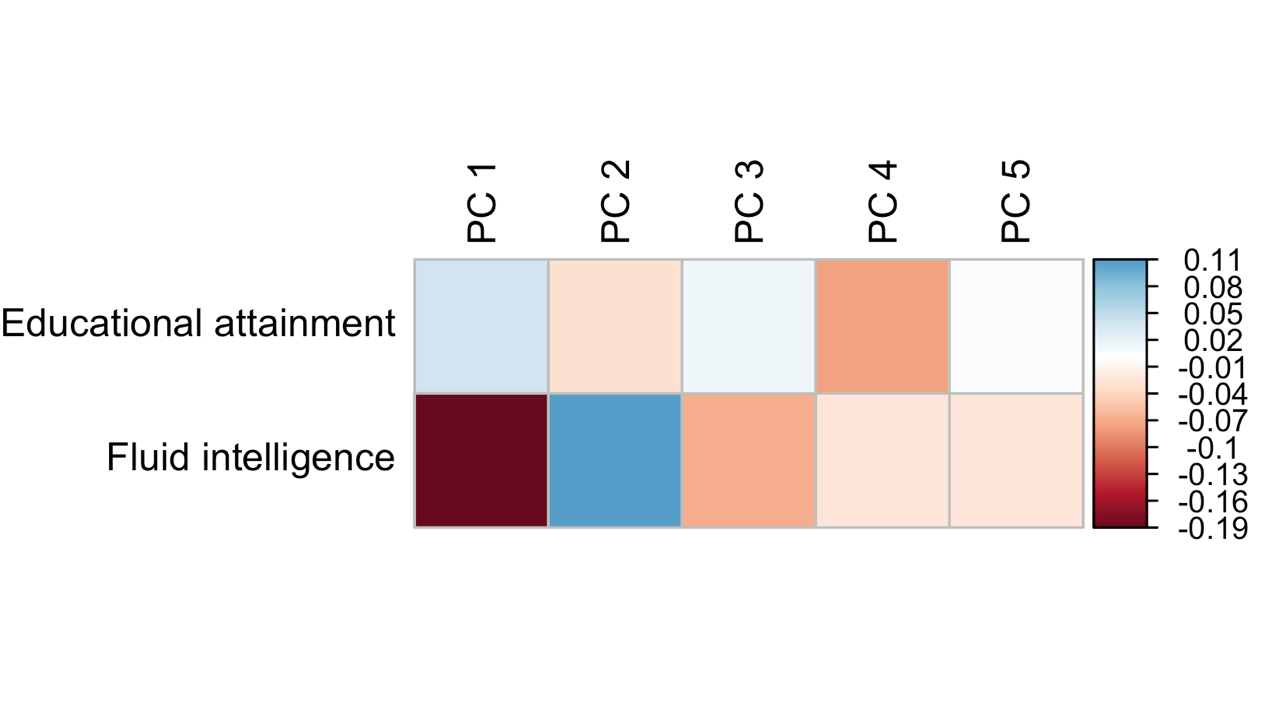

**Supplementary Figure 6.** Heatmap depicting the magnitude and significance of correlations between educational attainment and fluid intelligence (y-axis) and the top five genome-wide principal components (PCs) related to ancestry (x-axis) in the UK Biobank. The color of each box represented the magnitude and direction of the Pearson’s *r* correlation coefficient. All associations were statistically significant.

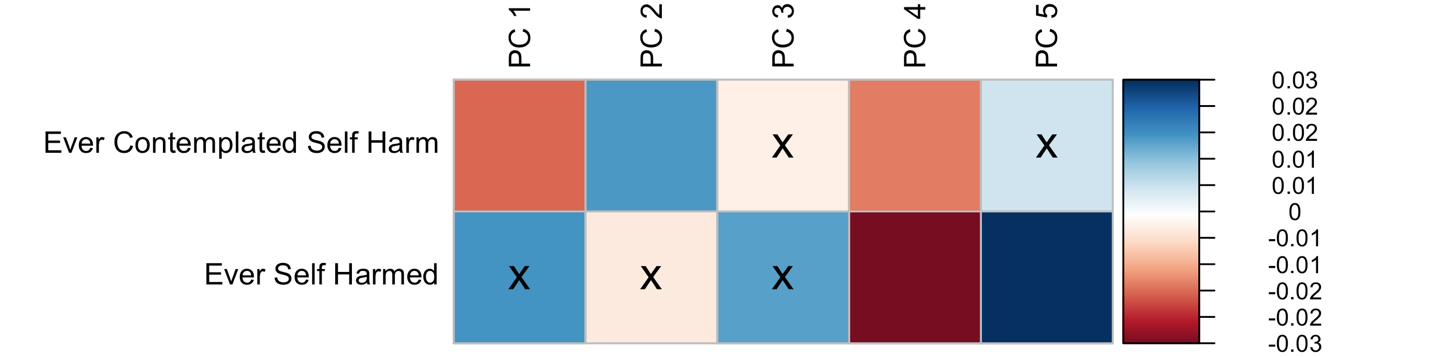

**Supplementary Figure 7.** Heatmap depicting the magnitude and significance of correlations between suicidality (y-axis) and the top five genome-wide principal components (PCs) related to ancestry (x-axis) in the UK Biobank. The color of each box represented the magnitude and direction of the Pearson’s *r* correlation coefficient. Boxes with no cross mark (“x”) represent nominally significant associations between suicidality and genome-wide PCs.

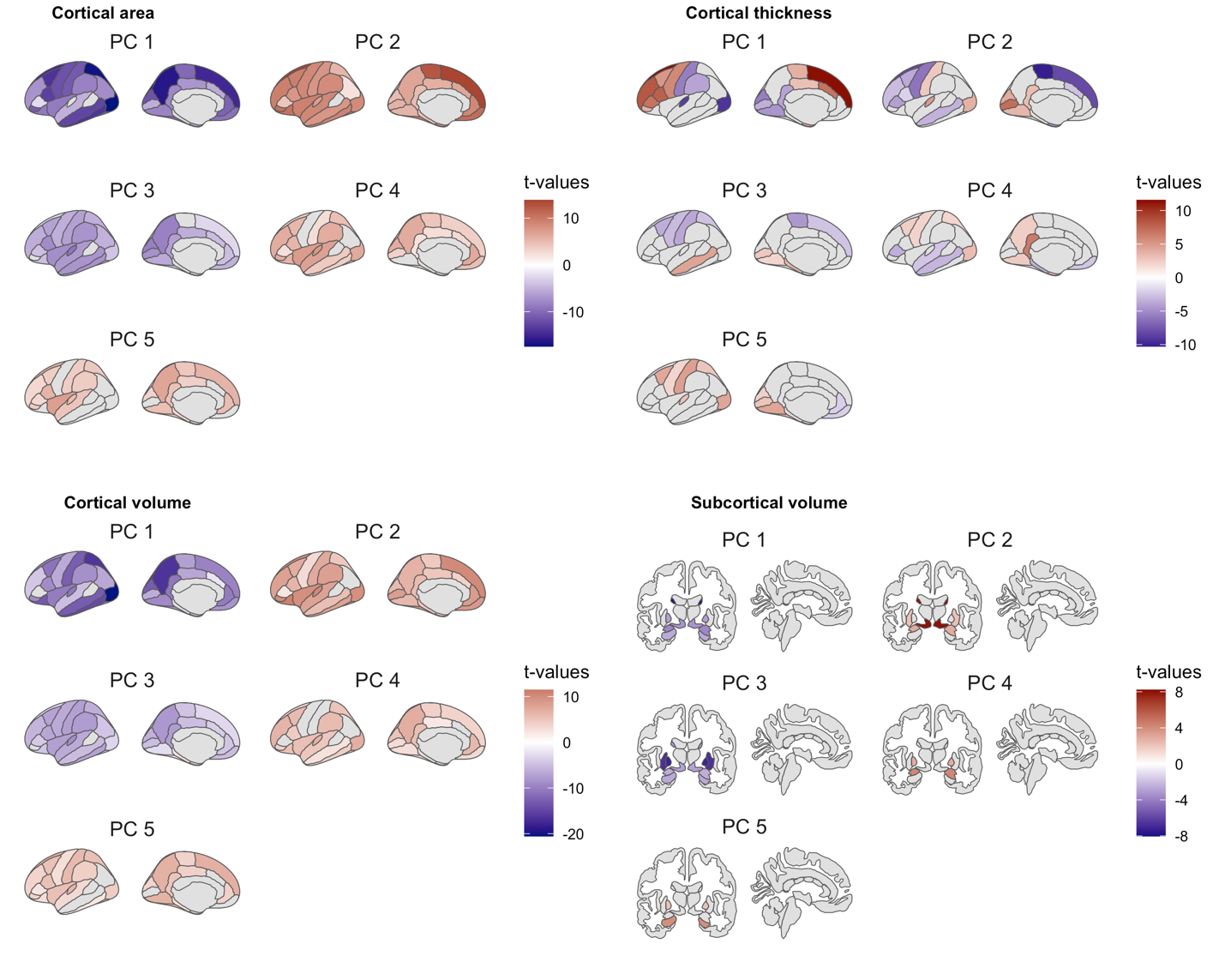

**Supplementary Figure 8**. Anatomical brain heatmap depicting brain regions with imaging-derived phenotypes (IDPs) that were significantly associated with the top five genome-wide principal components (PCs) related to ancestry in the UK Biobank. Brain regions are shaded according to the magnitude and direction of the *t*-statistic. All regions appearing in color (non-grey) showed at least a nominally significant association (uncorrected *p*<0.05) with one or more PCs. Regions in grey were not significantly associated with a PC.

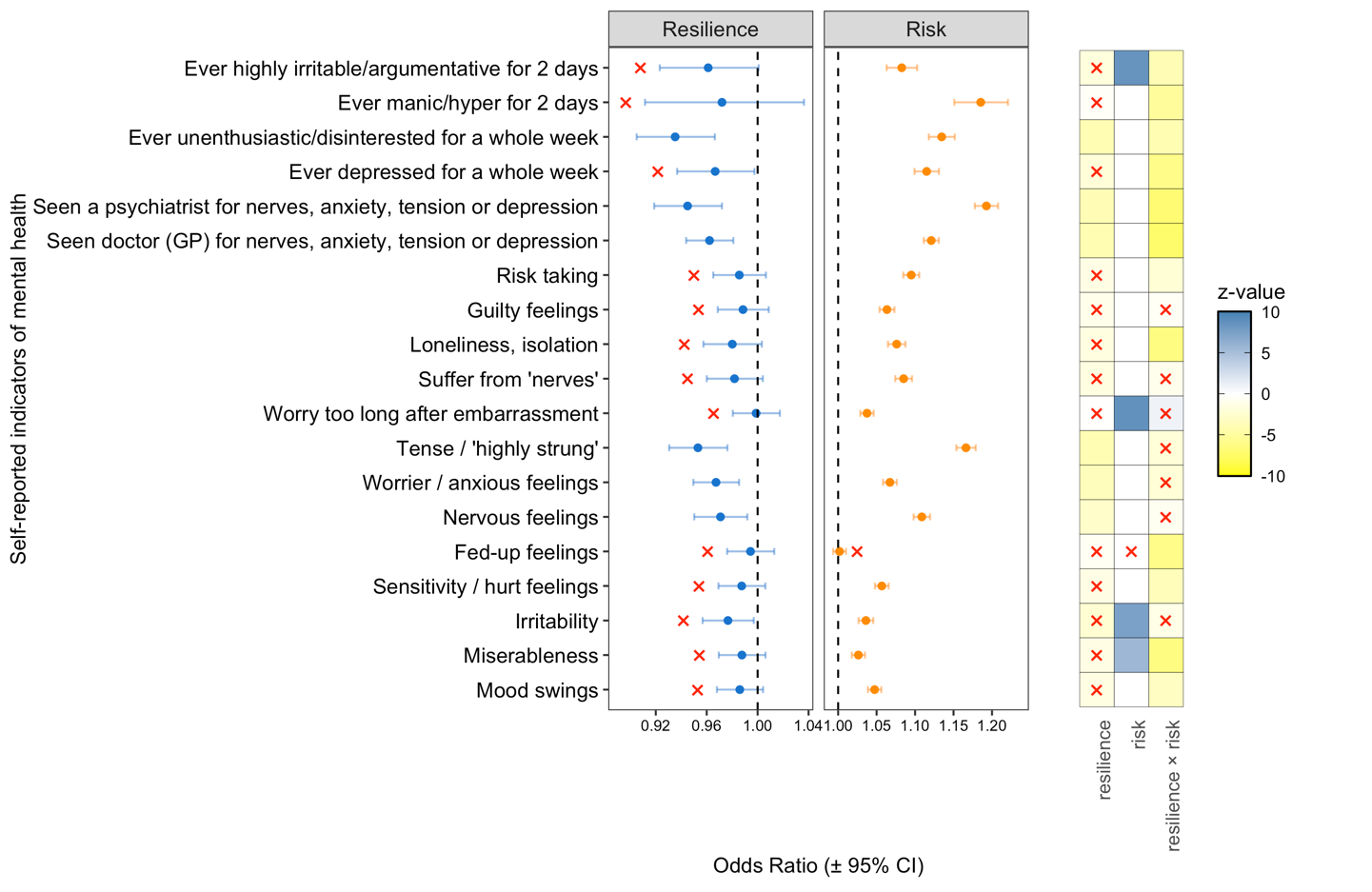

**Supplementary Figure 9**. Resilience scores moderate the penetrance of schizophrenia risk scores, enhancing self-reported mental well-being and reducing self-harm behaviors. A graphical representation of main and interactive effects of schizophrenia (SCZ) risk and resilience scores on 19 self-reported indicators of mental health provided by UK Biobank participants at the time of enrollment. All measures included in our analysis were scored as binary (yes/no) events. The results shown in this plot for resilience scores correspond to the scores calculated using resilience-associated SNPs that exhibit minimal linkage disequilibrium with mild-risk SNPs for SCZ (*R*^2^ value of 0.2 or less). The dot-plot on the left depicts the main effects of resilience (blue dots) and SCZ risk scores (orange dots), excluding their interactive effect, for each of the 19 corresponding items from the self-reported mental health questionnaire (y-axis). The main effects of resilience and SCZ risk are presented in terms of the Odds Ratio on the x-axis. The 95% confidence intervals of the Odds Ratio are shown as error bars in the bar-plot. Non-significant main effects (FDR*p* > 0.05) are denoted by a red “x”. The heat-map shown on the right side of the plot presents the significance (FDR*-*adjusted *p*-values, -log_10_ transformed) of the main effects of resilience and SCZ risk scores in the presence of their interactive effect (to enhance clarity, the range has been capped at -10 and +10). Similar to the bar-plot, non-significant main or interactive effects are denoted by a red “x”. A vertical dotted line is provided as a reference to an Odds Ratio of 1.0, a null effect, in each panel of the dot-plot.
